# Social Cognition in Temporal and Frontal Lobe Epilepsy: Systematic review, Meta-analysis, and Clinical Recommendations

**DOI:** 10.1101/2021.04.28.21255765

**Authors:** Maryam Ziaei, Charlotte Arnold, Kate Thompson, David Reutens

**Author notes:** Corresponding Author: Maryam Ziaei; Kavli Institute for Systems Neuroscience, Norwegian University of Science and Technology, Trondheim, Norway; and.

## Abstract

**Objective:** Despite the importance of social cognitive functions to mental health and social adjustment, assessment of these functions is absent in routine assessment of epilepsy patients. Thus, this review aims to provide a comprehensive overview of the literature on four major aspects of social cognition among temporal and frontal lobe epilepsy, which is a critical step towards designing new interventions.

**Method:** Papers from 1990-2021 were reviewed and examined for inclusion in this study. After the deduplication process, a systematic review and meta-analysis of 44 and 40 articles, respectively, involving 113 people with frontal lobe epilepsy and 1482 people with temporal lobe epilepsy were conducted.

**Results:** Our results indicated that while patients with frontal or temporal lobe epilepsy have difficulties in all aspects of social cognition relative to non-clinical controls, the effect sizes were larger for theory of mind (*g*= 0.95), than for emotion recognition (*g*=0.69) among temporal lobe epilepsy group. The frontal lobe epilepsy group exhibited significantly greater impairment in emotion recognition compared to temporal lobe. Additionally, people with right temporal lobe epilepsy (*g*= 1.10) performed more poorly than those with a left-sided (*g*= 0.90) seizure focus, specifically in the theory of mind domain.

**Conclusions:** These data point to a potentially important difference in the severity of deficits within the emotion recognition and theory of mind abilities depending on the laterlization of seizure side. We also suggest a guide for the assessment of impairments in social cognition that can be integrated into multidisciplinary clinical evaluation for people with epilepsy

## Introduction

Social cognition broadly refers to one’s ability to perceive and understand other people’s thoughts, feelings, and behaviors and to respond appropriately. The most researched aspects of social cognition are emotion recognition, theory of mind (ToM) and empathy. Emotion recognition is the ability to identify and discriminate emotional states from verbal and nonverbal cues. Theory of mind refers to the ability to understand what other people are thinking and feeling and to infer complex mental states, such as intention and disposition, in others. Empathy is the ability to understand and respond to the emotional experiences of others and has been shown to contribute to successful social interaction and to promote pro-social actions (Sun et al., 2019). The literature typically differentiates between two components of empathy: affective empathy, which entails affective sharing of other people’s emotional states, and cognitive empathy, which refers to the ability to decode and understand other people’s perspective (Singer & Lamm, 2009).

Social cognition is complex and multifaceted, subserved by an intricate network of interconnected brain regions collectively referred to as the “social brain”. These regions include the amygdala, medial prefrontal cortex, temporoparietal junction, anterior cingulate and insula. Many of these structures are adversely affected in conditions such as schizophrenia (Green et al., 2015), stroke (Hillis, 2014), and neurodegenerative disorders (Christidi et al., 2018), giving rise to the propensity for social cognitive impairment in these groups. Social cognitive impairment is a feature of many developmental, psychiatric, neurological and neurodegenerative conditions (Cotter et al., 2018). Not surprising, people with epilepsy also demonstrate impairments in social cognition (for instance, Giovagnoli et al., 2011; Giovagnoli et al., 2013) that negatively impact the quality of life, employability, and other cognitive functions. Despite the increasing volume of studies investigating social cognition in epilepsy over recent years, research into the predictors of social cognitive impairment in this population remains lacking. Specifically, the extent to which social cognitive difficulties are caused by medication, psychological, and social factors (e.g., fear of seizures, perceived stigma, discrimination, lack of social support), recurrent seizures, or the underlying epileptogenic brain lesion, remains unclear. Identifying the nature and magnitude of social cognitive impairments in people with epilepsy has both theoretical and clinical implications, including the potential to inform guidelines for clinical assessment and psychosocial intervention.

Four prior meta-analyses evaluated theory of mind and emotion recognition in patients with epilepsy. Outcomes differed slightly depending on the groups of patients studied and the outcome measures used. The chief findings of the studies were: (a) theory of mind ability was affected in patients with frontal lobe epilepsy (FLE) and temporal lobe epilepsy (TLE), but not from seizure disorders originating outside these areas (extra-TLE/FLE; Stewart et al., 2016); (b) deficient recognition of fear, followed by sadness and disgust, in both visual and auditory modalities was reported in patients with epilepsy (Edwards et al., 2017; Monti et al., 2015); (c) patients with right TLE exhibited more significant impairments in recognition of fear, disgust, and sadness than patient with left TLE (Bora et al., 2016); and (d) patients undergoing surgery did not differ in social cognitive outcomes (emotion recognition and theory of mind) compared to those not undergoing surgery (Bora et al., 2016).

While these previous meta-analytic reviews are a valuable contribution to the literature, it is still unknown how empathy and social behavior is affected by epilepsy. This study is the first to evaluate all four major components of social cognition, emotion recognition, theory of mind, empathy and social behavior, in people with TLE and FLE. We acknowledge that social cognition is a multidimensional construct and significantly influenced by personality vulnerabilities, mood disturbance and cognitive impairment. However, for the purpose of this review, we examine the evidence for four social cognitive components previously investigated in other clinical populations (Henry et al., 2016; Adams et al., 2019). Our primary aims were twofold: (a) to determine whether social cognitive impairment is global or specific to one domain of social cognition in people with TLE and FLE, and (b) to investigate whether the severity and patterns of impairment are moderated by the location of the epileptic focus in the temporal vs. the frontal lobe. We conclude this review with a suggested pathway and clinically useful tools to support the screening of social cognition in people with epilepsy when indicators of potential compromise are identified.

## Method

This systematic review was conducted in line with the Preferred Reporting Items for Systematic Review and Meta-Analyses (PRISMA) guidelines (Moher et al., 2009). The completed PRISMA checklist is presented in the supplementary material.

### Identification of studies

#### Search strategy

We searched several databases from 1990 to January 21 including PsycINFO and MEDLINE (via EBSCOhost), Embase, and Web of Science Core Collection (via Clarivate Analytics). We constructed a comprehensive search strategy using variant terms for the focal epilepsy subtypes and social cognition constructs. The search strategy developed for articles is presented below; the full search strategy can be made available upon request.

(“epilep*” OR “focal epilepsy” OR “temporal lobe epilepsy” OR “frontal lobe epilepsy” OR “seizure” OR ((“amygdala” OR “hippocamp*”) AND “damage”)) AND (“perc*” OR “identif*” OR “recogni*” OR “process*” OR “label*”) AND (“emotion recognition” OR ((“face” OR “facial”) AND (“affect*” OR “emotion*” OR “expression*”)) OR “theory of mind” OR “social cognition” OR “social perception” OR “perspective task*” OR “mentalis*” OR “mind read*” OR “empath*” OR “social competence” OR “social outcome*” OR “social adjustment” OR “social behavio*” OR “social skill*” OR “social interaction*”).

Duplicate results were removed automatically in EndNote. Using the Ancestry Method, reference lists were searched for appropriate reviews and eligible studies to identify additional papers. Then titles and abstracts were screened by two authors (MZ & CA). Full-text of papers were reviewed when it was unclear whether an article fulfilled the eligibility criteria based on the title and abstract alone. Two authors (MZ & CA) independently screened the full text articles against the inclusion criteria and discrepancies were resolved through discussion.

#### Inclusion criteria

The review included studies that: 1. reported primary research published in English-language peer-reviewed journals; 2. included samples of people diagnosed with temporal lobe (TLE) or frontal lobe epilepsy (FLE) and reported the data separately for each patient group; 3. included a non-clinical comparison group; 4. used validated measures evaluating each of the four components of social cognition (as detailed in Table 1); and 5. included either paediatric or adult patient populations.

**Table 1.**
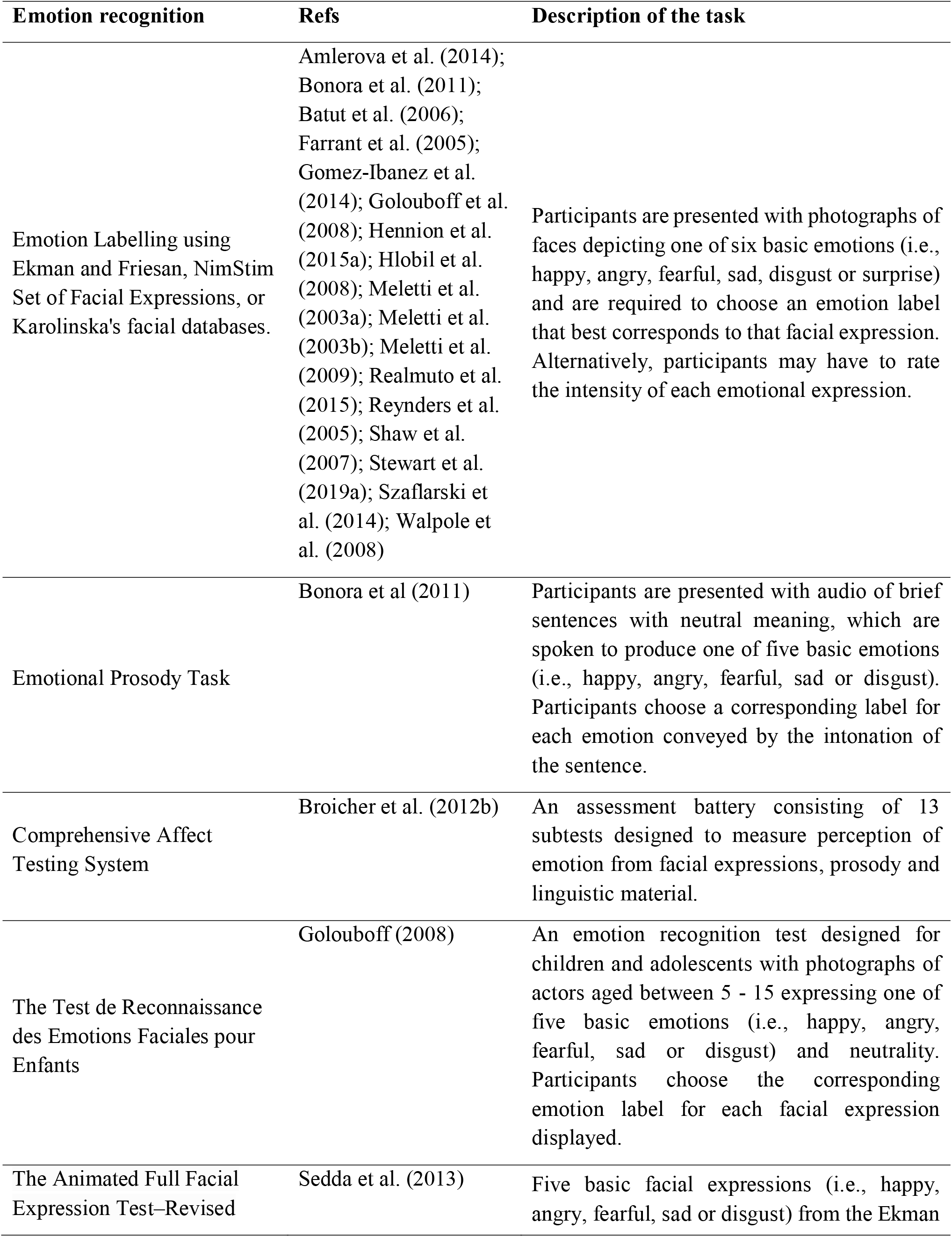
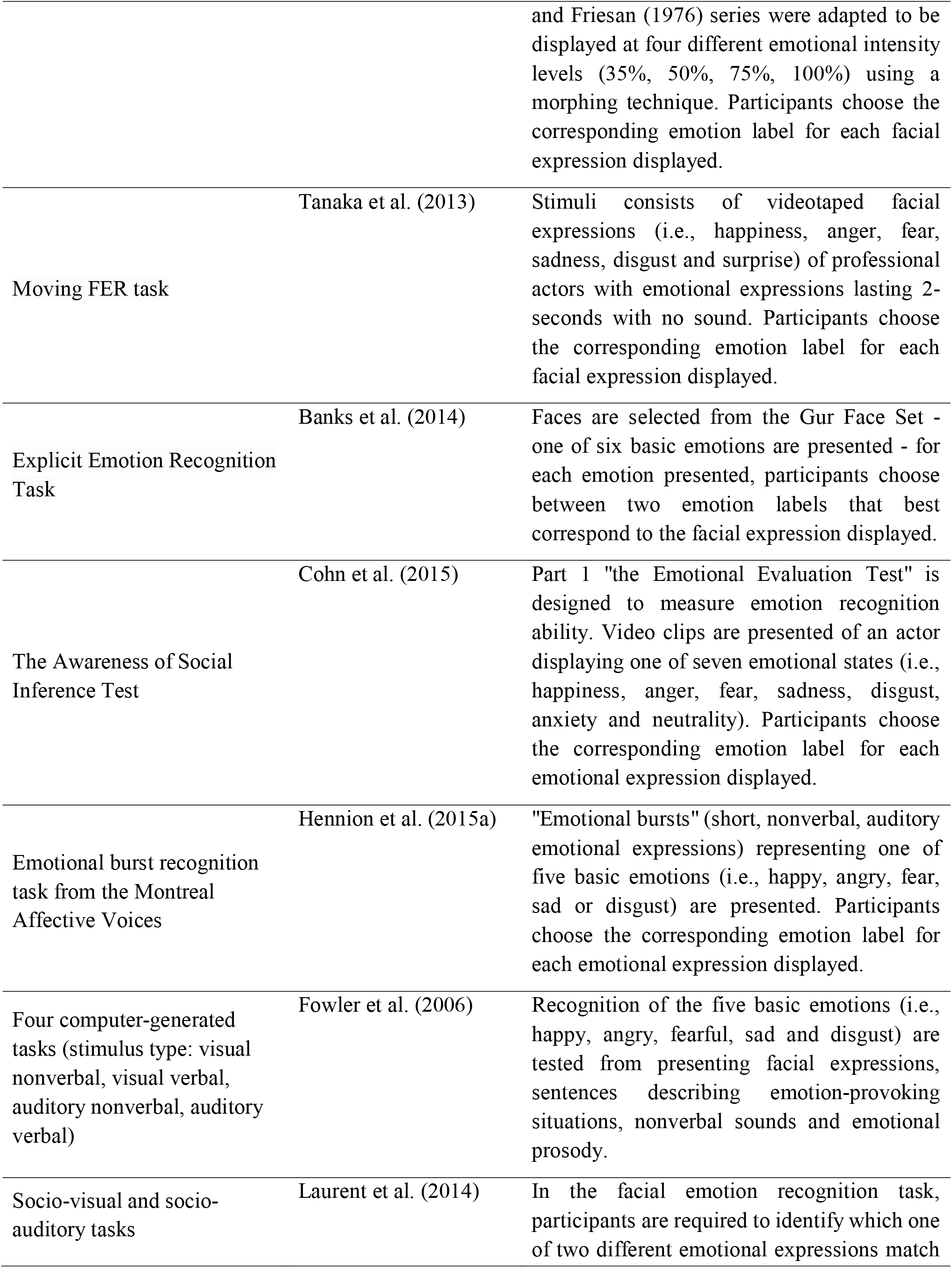
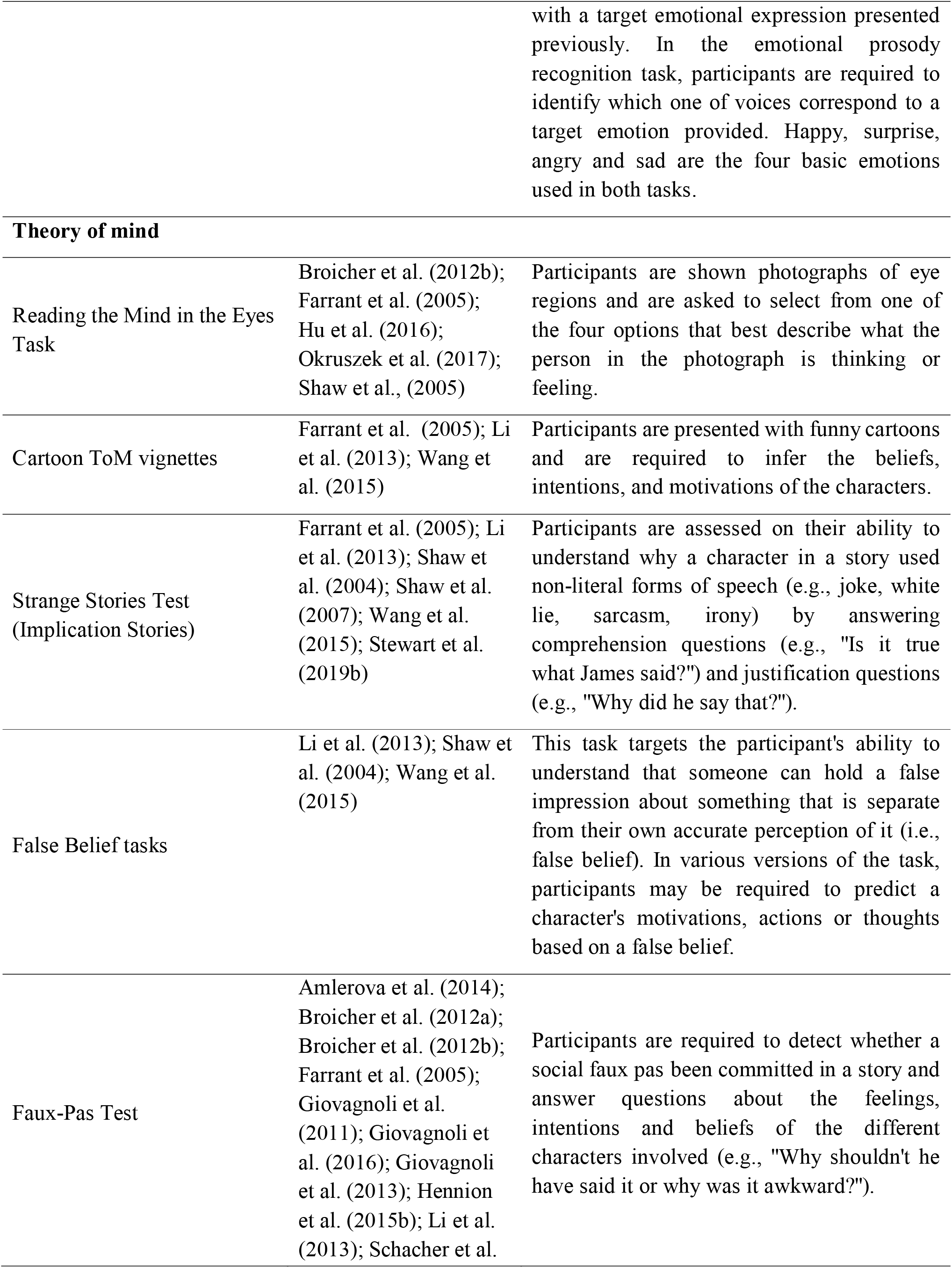
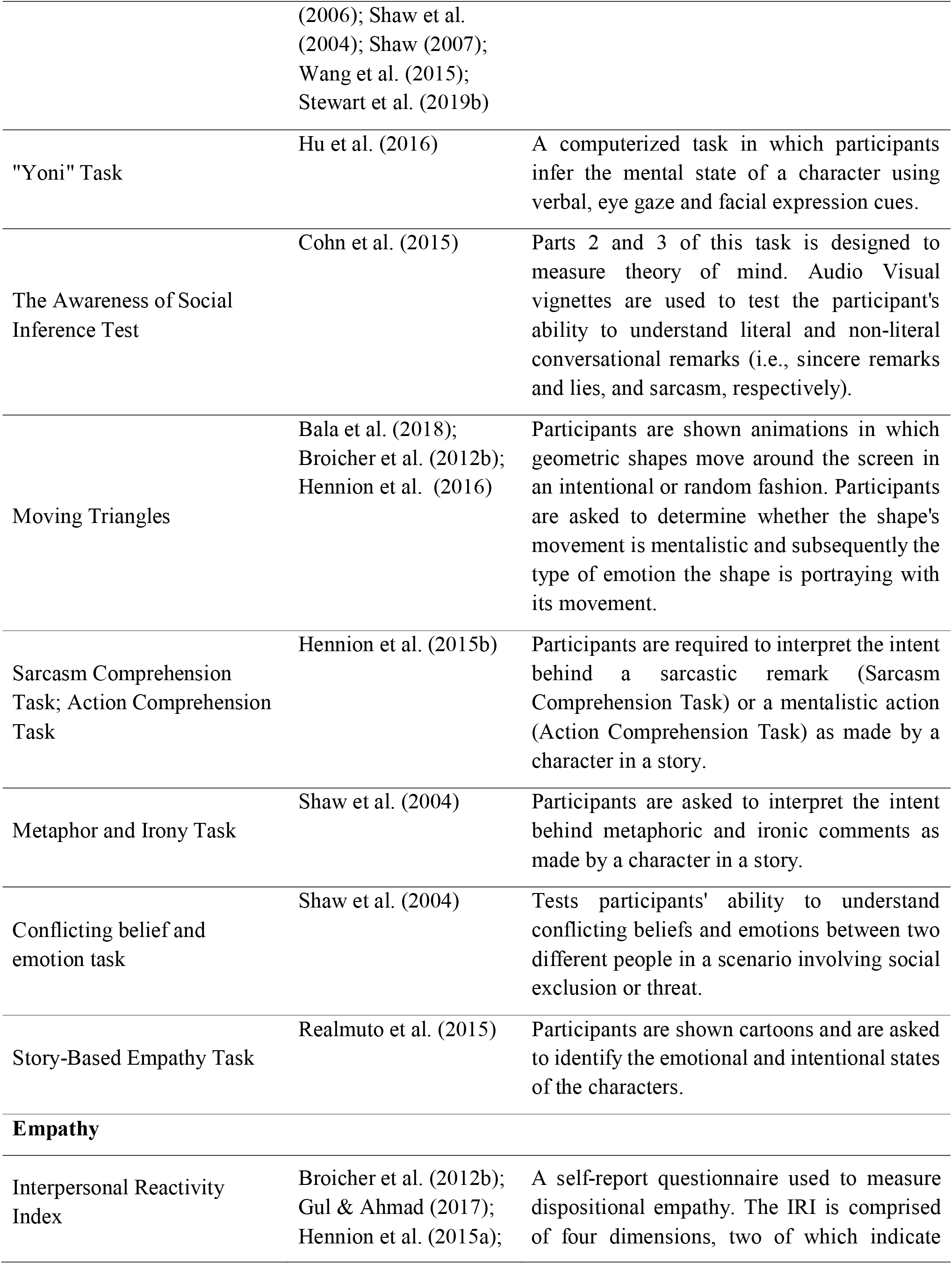
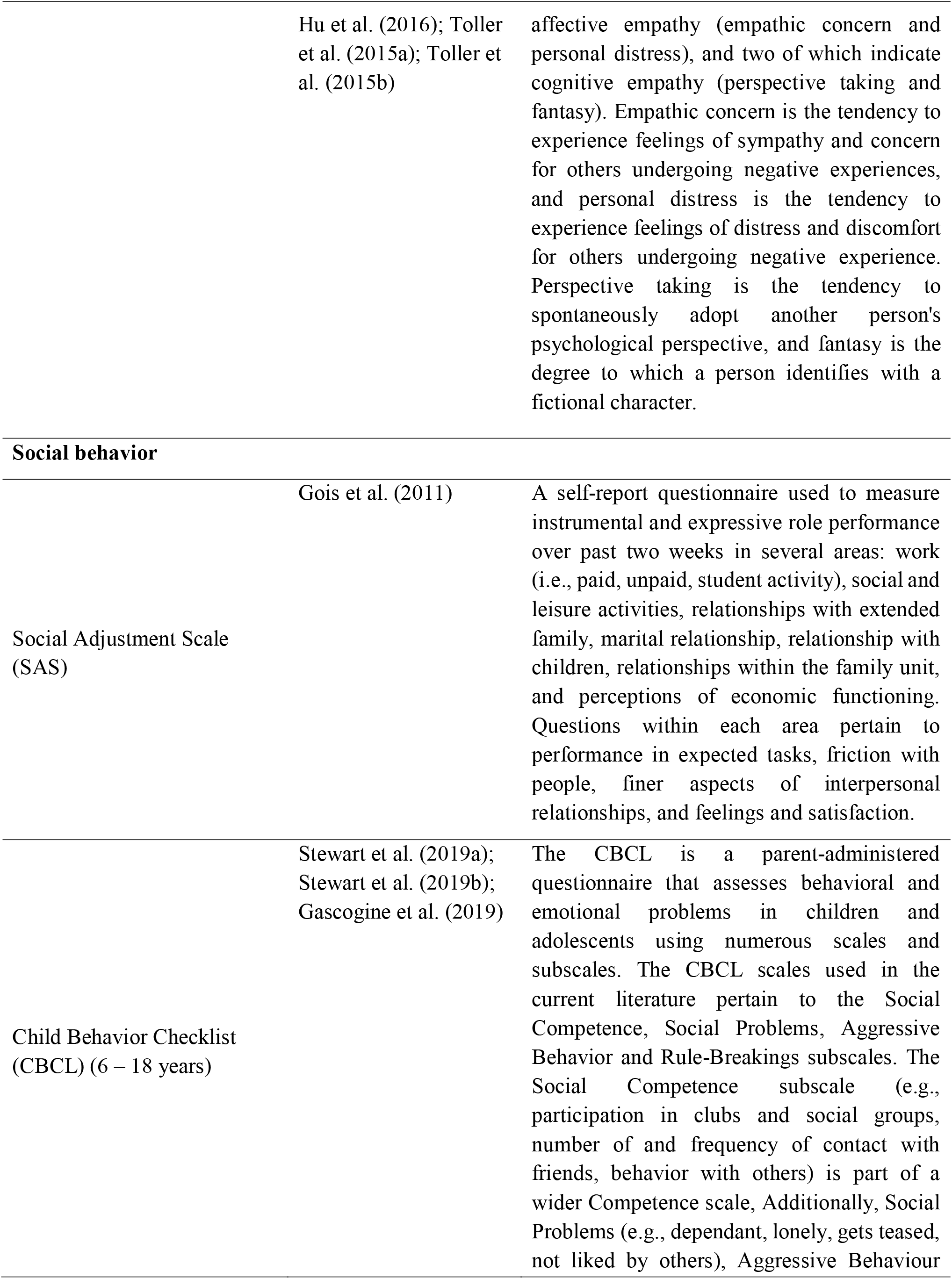
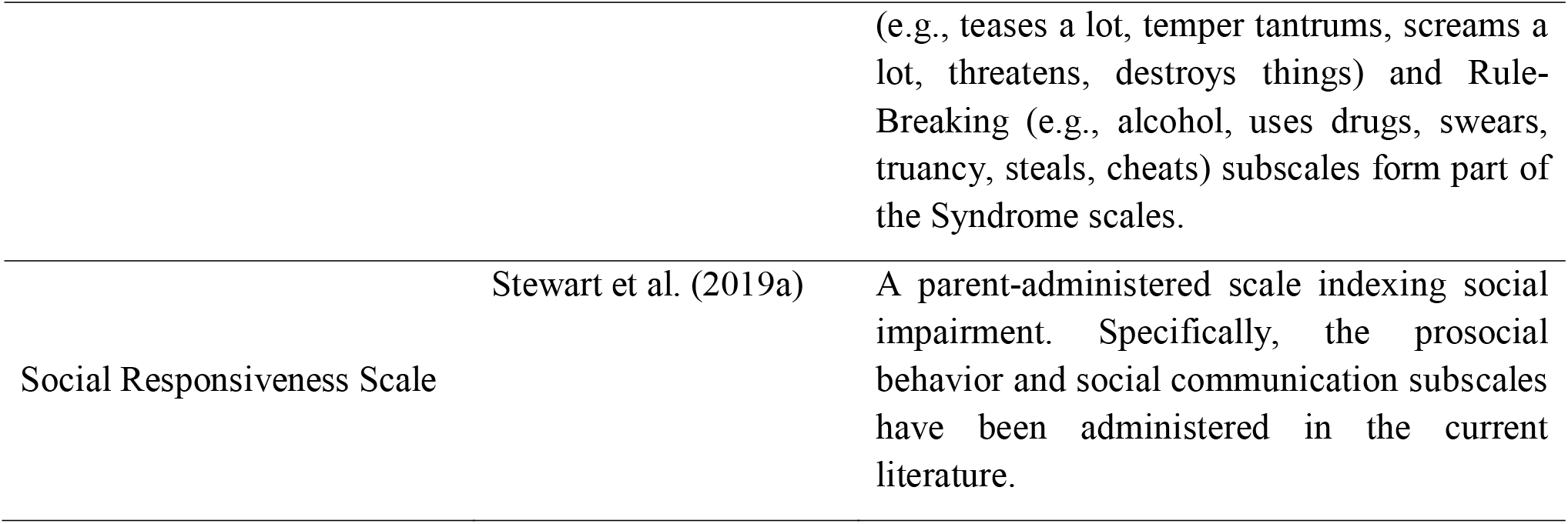
Description of measures used for each social cognitive function.

#### Exclusion criteria

The review excluded: 1. case studies, non-clinical outcome studies (e.g., psychometric validation studies), secondary research or special papers (e.g., reviews, editorial, commentaries, or letters); 2. studies that did not include a control group; 3. studies that recruited heterogenous focal epilepsy groups (e.g., multifocal epilepsy from frontal and temporal lobes); 4. studies that used qualitative measures such as interviews or behavioral tasks other than those specified in Table 1; 6. studies with inadequate data to calculate a mean or weighted effect; 7. fMRI studies that investigated one of these domains in TLE or FLE patients but without overt, explicit, behavioral measures during the fMRI task; 8. studies that included patients with unilateral or bilateral amygdala or hippocampal damage, but without seizures; 9. studies that included only post-operative patients, or grouped patients with and without lobectomy together. Studies were not excluded based on other premorbid conditions, cause of epilepsy (e.g., acquired lesions, tumours, congenital structural abnormalities), or medication use.

### Definition of social cognitive domains

Definition of each domain is extensively discussed in the supplementary materials. In short, in the following we provide a brief description of each domain:

*Emotion recognition*: we defined emotion recognition as any task that required participants to label, recognize, rate, match or select the emotions expressed within the stimuli. Stimuli could be visual or auditory and may consist of static faces, videos, or sounds. All eligible studies evaluating emotion recognition and their effect sizes are reported in Table 2.

**Table 2.**
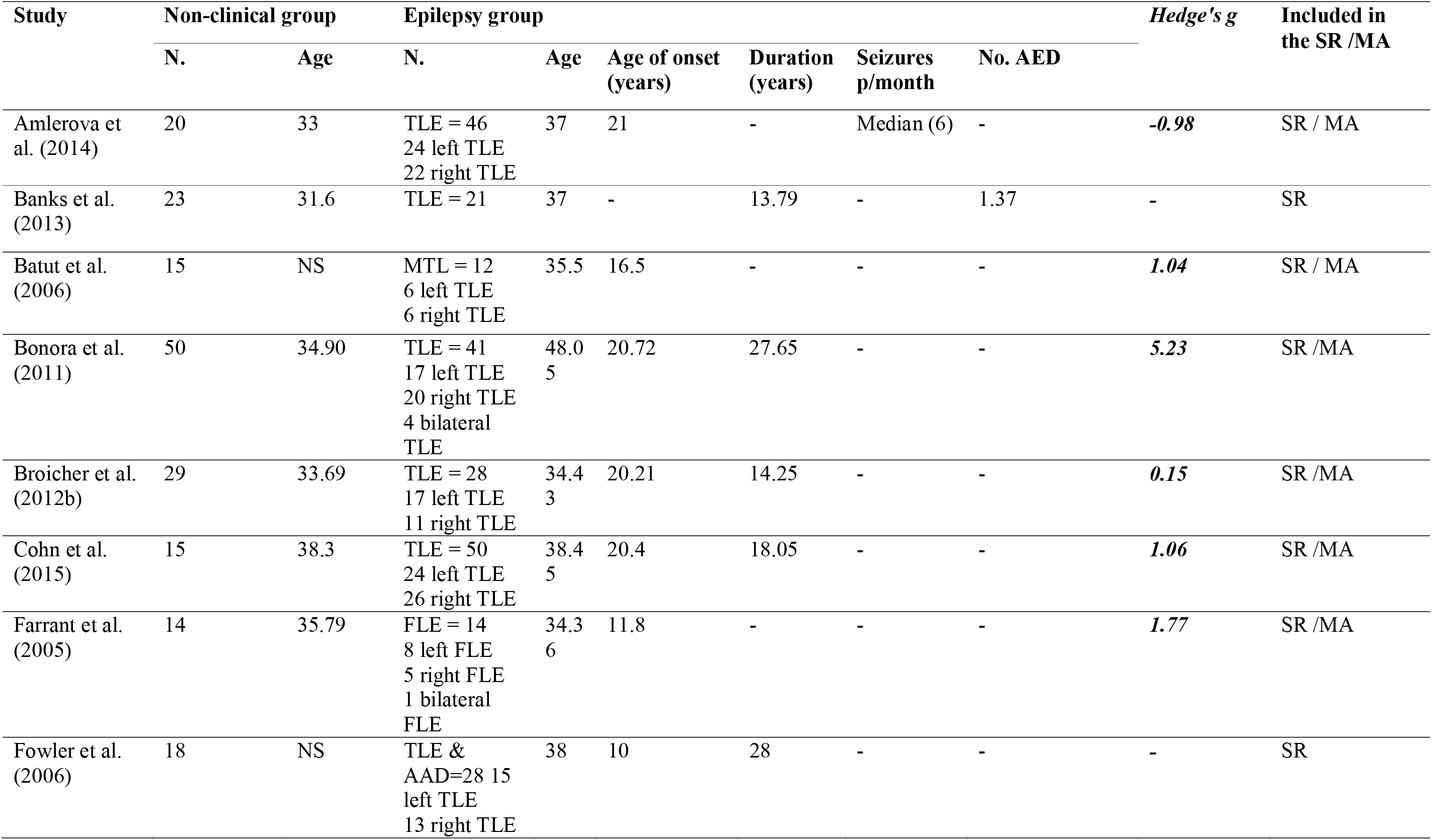
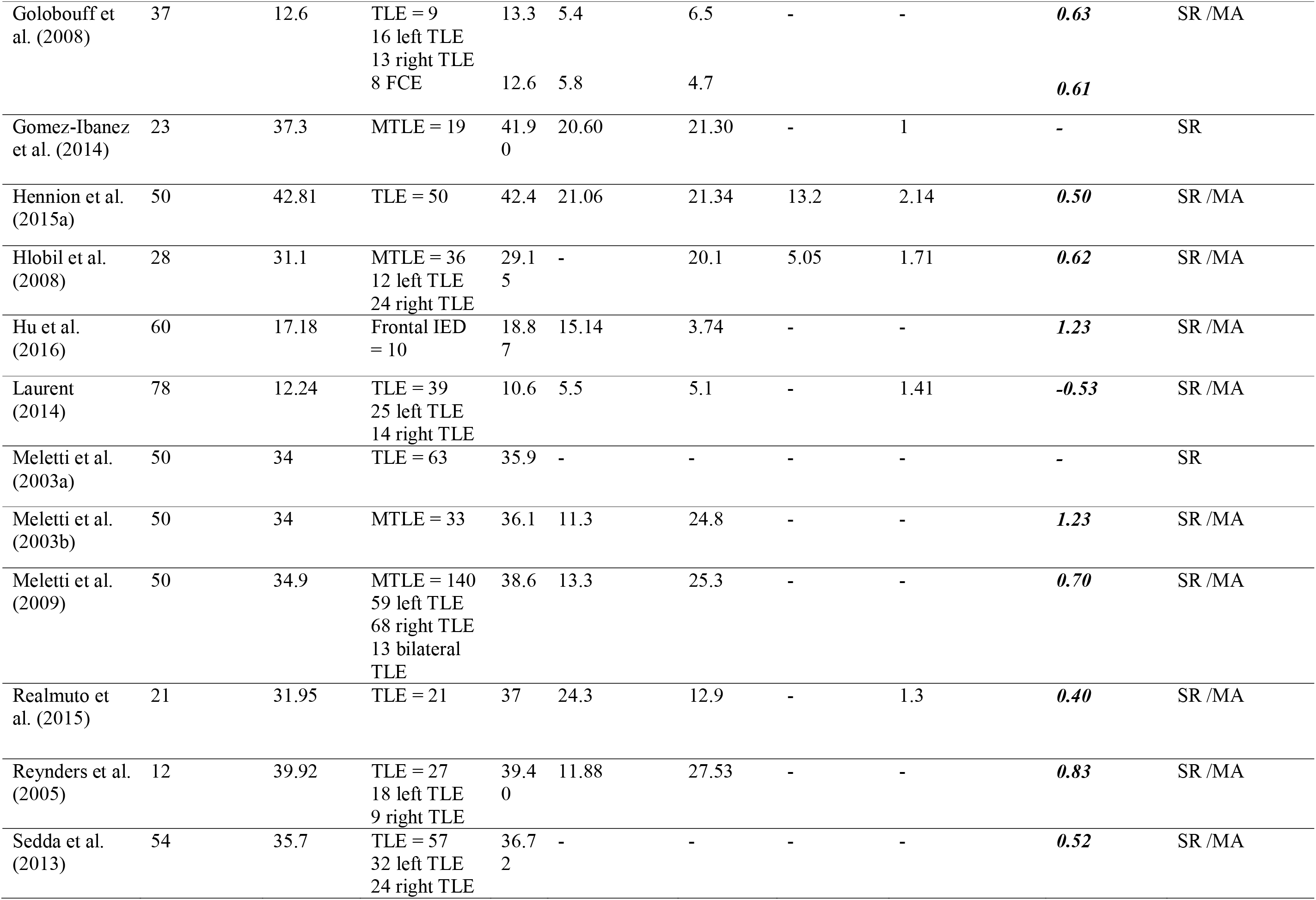
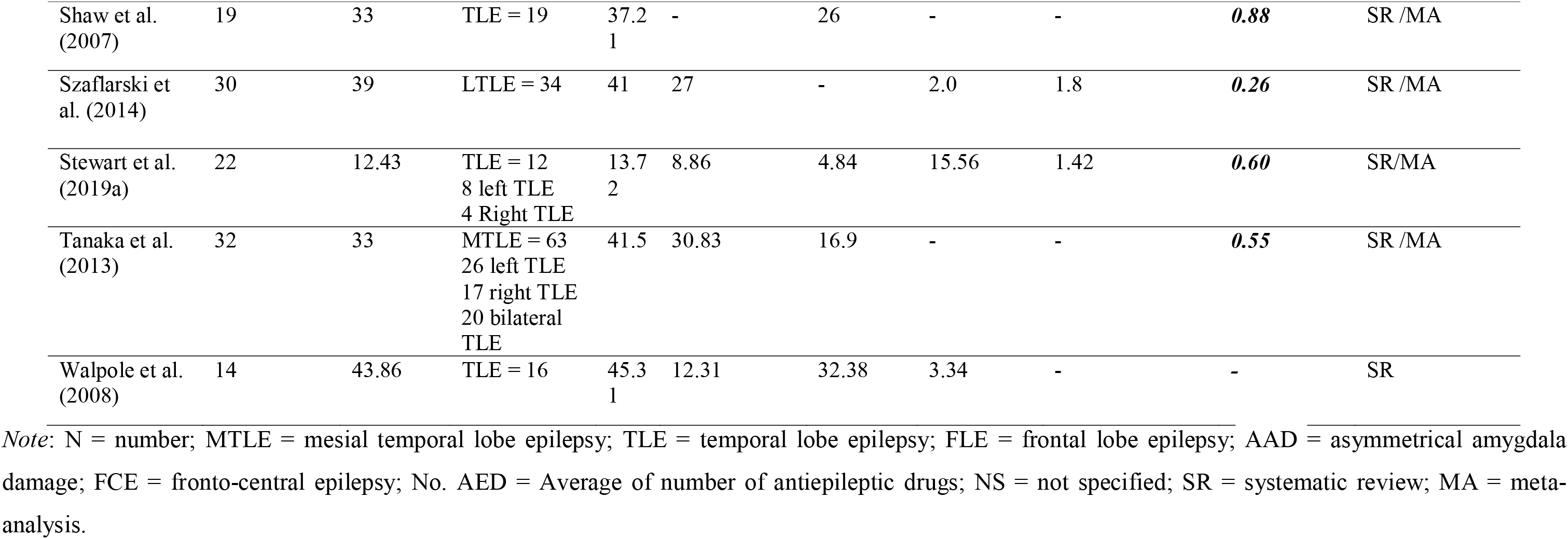
Study characteristics and effect sizes (Hedge’s g) reported individually for studies included in the meta-analysis pertaining to emotion perception.

| Study | Non-clinical group |  | Epilepsy group |  |  |  |  |  | Hedge's *g* | Included in the SR /MA |
| --- | --- | --- | --- | --- | --- | --- | --- | --- | --- | --- |
|  | N. | Age | N. | Age | Age of onset (years) | Duration (years) | Seizures p/month | No. AED |  |  |
| Amlerova et al. (2014) | 20 | 33 | TLE = 46<br>24 left TLE<br>22 right TLE | 37 | 21 | - | Median (6) | - | **-0.98** | SR / MA |
| Banks et al. (2013) | 23 | 31.6 | TLE = 21 | 37 | - | 13.79 | - | 1.37 | **-** | SR |
| Batut et al. (2006) | 15 | NS | MTL = 12<br>6 left TLE<br>6 right TLE | 35.5 | 16.5 | - | - | - | **1.04** | SR / MA |
| Bonora et al. (2011) | 50 | 34.90 | TLE = 41<br>17 left TLE<br>20 right TLE<br>4 bilateral TLE | 48.0<br>5 | 20.72 | 27.65 | - | - | **5.23** | SR /MA |
| Broicher et al. (2012b) | 29 | 33.69 | TLE = 28<br>17 left TLE<br>11 right TLE | 34.4<br>3 | 20.21 | 14.25 | - | - | **0.15** | SR /MA |
| Cohn et al. (2015) | 15 | 38.3 | TLE = 50<br>24 left TLE<br>26 right TLE | 38.4<br>5 | 20.4 | 18.05 | - | - | **1.06** | SR /MA |
| Farrant et al. (2005) | 14 | 35.79 | FLE = 14<br>8 left FLE<br>5 right FLE<br>1 bilateral FLE | 34.3<br>6 | 11.8 | - | - | - | **1.77** | SR /MA |
| Fowler et al. (2006) | 18 | NS | TLE & AAD=28<br>15 left TLE<br>13 right TLE | 38 | 10 | 28 | - | - | **-** | SR |
| Golobouff et al. (2008) | 37 | 12.6 | TLE = 9<br>16 left TLE<br>13 right TLE<br>8 FCE | 13.3<br>12.6 | 5.4<br>5.8 | 6.5<br>4.7 | - | - | **0.63**<br>**0.61** | SR /MA |
| Gomez-Ibanez et al. (2014) | 23 | 37.3 | MTLE = 19 | 41.9<br>0 | 20.60 | 21.30 | - | 1 | - | SR |
| Hennion et al. (2015a) | 50 | 42.81 | TLE = 50 | 42.4 | 21.06 | 21.34 | 13.2 | 2.14 | **0.50** | SR /MA |
| Hlobil et al. (2008) | 28 | 31.1 | MTLE = 36<br>12 left TLE<br>24 right TLE | 29.1<br>5 | - | 20.1 | 5.05 | 1.71 | **0.62** | SR /MA |
| Hu et al. (2016) | 60 | 17.18 | Frontal IED = 10 | 18.8<br>7 | 15.14 | 3.74 | - | - | **1.23** | SR /MA |
| Laurent (2014) | 78 | 12.24 | TLE = 39<br>25 left TLE<br>14 right TLE | 10.6 | 5.5 | 5.1 | - | 1.41 | **-0.53** | SR /MA |
| Meletti et al. (2003a) | 50 | 34 | TLE = 63 | 35.9 | - | - | - | - | - | SR |
| Meletti et al. (2003b) | 50 | 34 | MTLE = 33 | 36.1 | 11.3 | 24.8 | - | - | **1.23** | SR /MA |
| Meletti et al. (2009) | 50 | 34.9 | MTLE = 140<br>59 left TLE<br>68 right TLE<br>13 bilateral TLE | 38.6 | 13.3 | 25.3 | - | - | **0.70** | SR /MA |
| Realmuto et al. (2015) | 21 | 31.95 | TLE = 21 | 37 | 24.3 | 12.9 | - | 1.3 | **0.40** | SR /MA |
| Reynders et al. (2005) | 12 | 39.92 | TLE = 27<br>18 left TLE<br>9 right TLE | 39.4<br>0 | 11.88 | 27.53 | - | - | **0.83** | SR /MA |
| Sedda et al. (2013) | 54 | 35.7 | TLE = 57<br>32 left TLE<br>24 right TLE | 36.7<br>2 | - | - | - | - | **0.52** | SR /MA |
| Shaw et al. (2007) | 19 | 33 | TLE = 19 | 37.2<br>1 | - | 26 | - | - | **0.88** | SR /MA |
| Szaflarski et al. (2014) | 30 | 39 | LTLE = 34 | 41 | 27 | - | 2.0 | 1.8 | **0.26** | SR /MA |
| Stewart et al. (2019a) | 22 | 12.43 | TLE = 12<br>8 left TLE<br>4 Right TLE | 13.7<br>2 | 8.86 | 4.84 | 15.56 | 1.42 | **0.60** | SR/MA |
| Tanaka et al. (2013) | 32 | 33 | MTLE = 63<br>26 left TLE<br>17 right TLE<br>20 bilateral TLE | 41.5 | 30.83 | 16.9 | - | - | **0.55** | SR /MA |
| Walpole et al. (2008) | 14 | 43.86 | TLE = 16 | 45.3<br>1 | 12.31 | 32.38 | 3.34 | - | - | SR |

*Note:* N = number; MTLE = mesial temporal lobe epilepsy; TLE = temporal lobe epilepsy; FLE = frontal lobe epilepsy; AAD = asymmetrical amygdala damage; FCE = fronto-central epilepsy; No. AED = Average of number of antiepileptic drugs; NS = not specified; SR = systematic review; MA = meta-analysis.

*Theory of mind*: we defined theory of mind as the ability to understand others’ thinking and feeling and included studies that used measures that were commonly used in the social cognitive literature such as False Belief, Faux Pas, or Reading the Mind in the Eyes Test. All eligible studies evaluating theory of mind and their effect sizes are reported in Table 3.

**Table 3.**
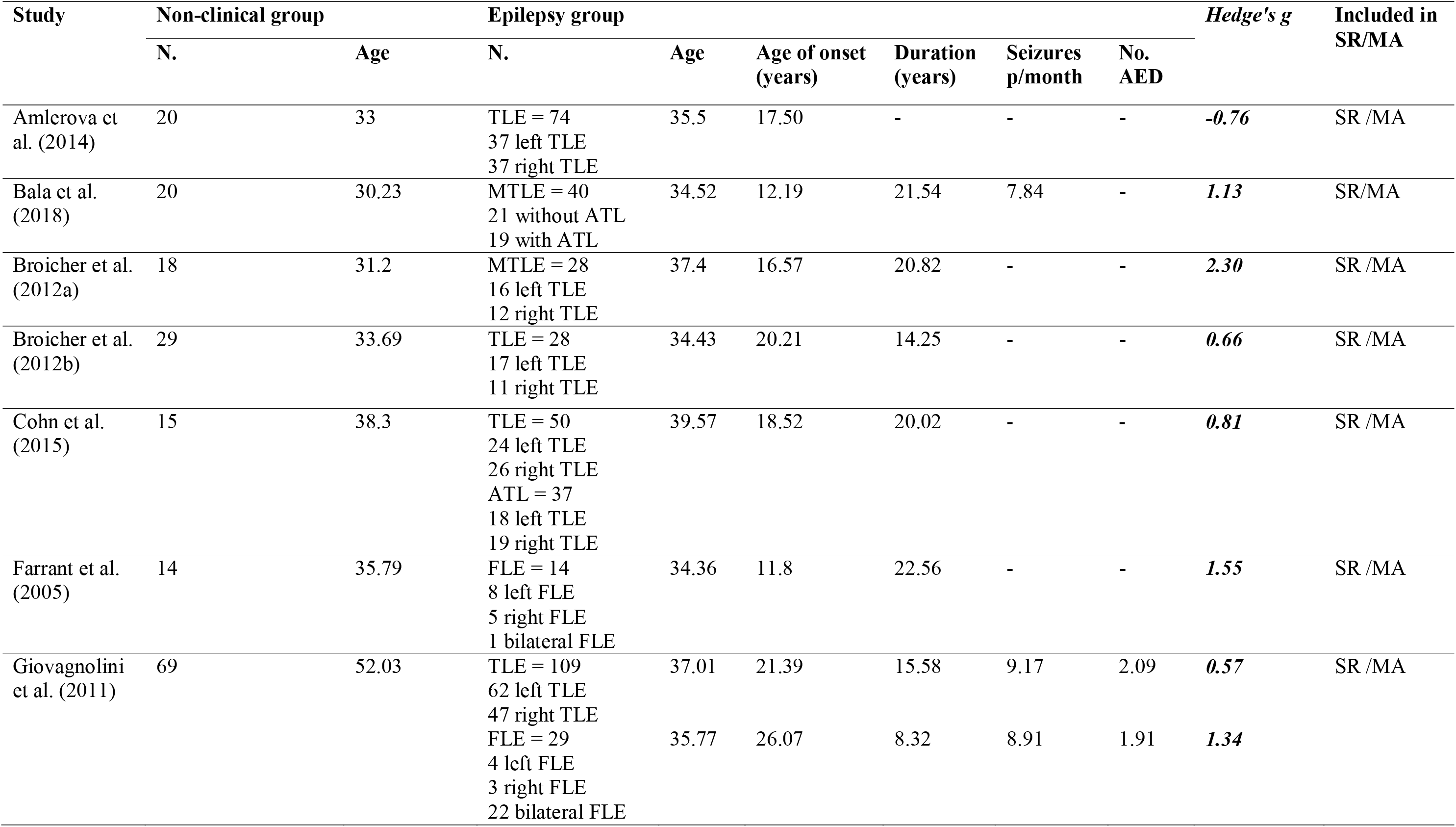
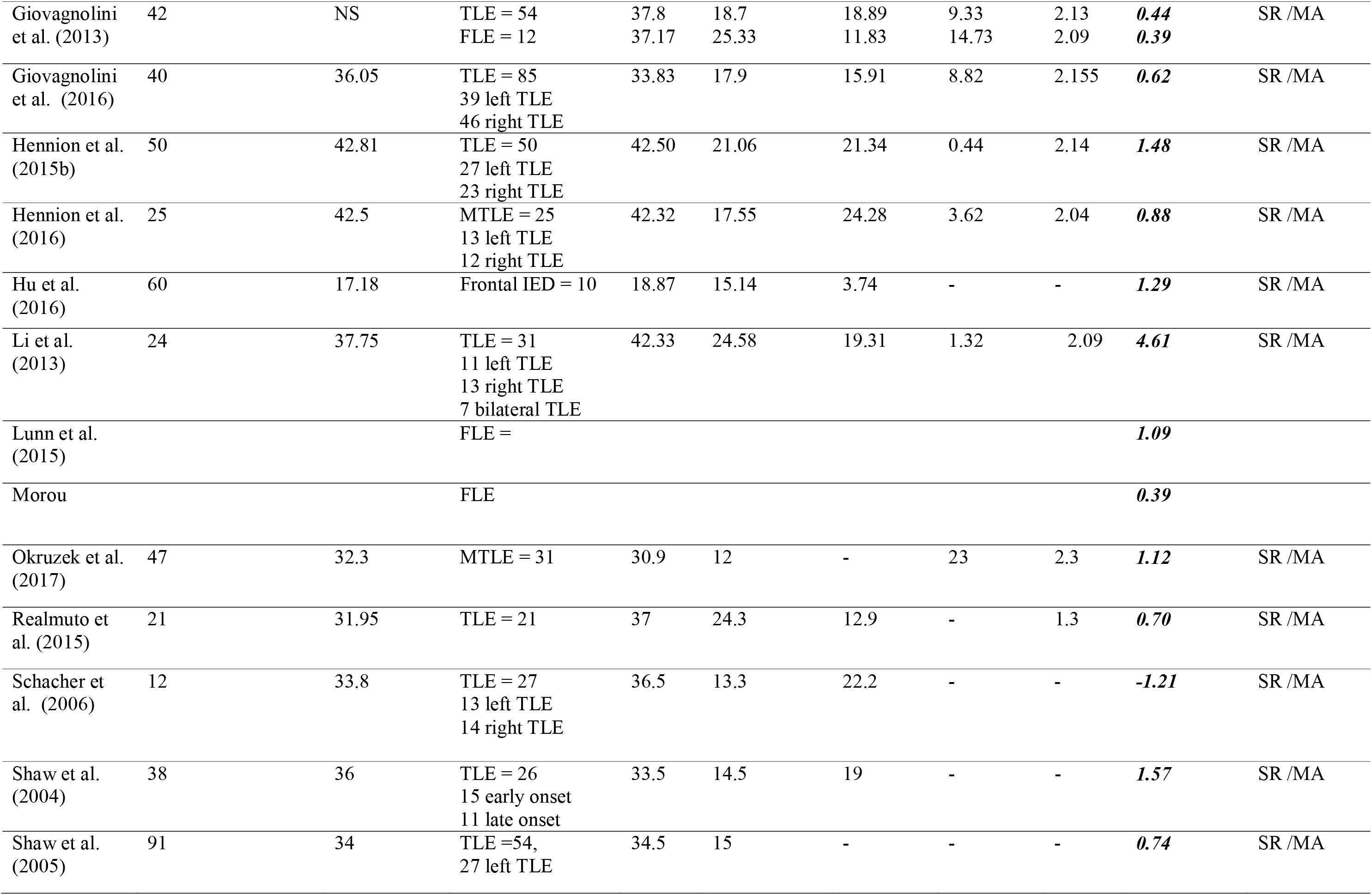
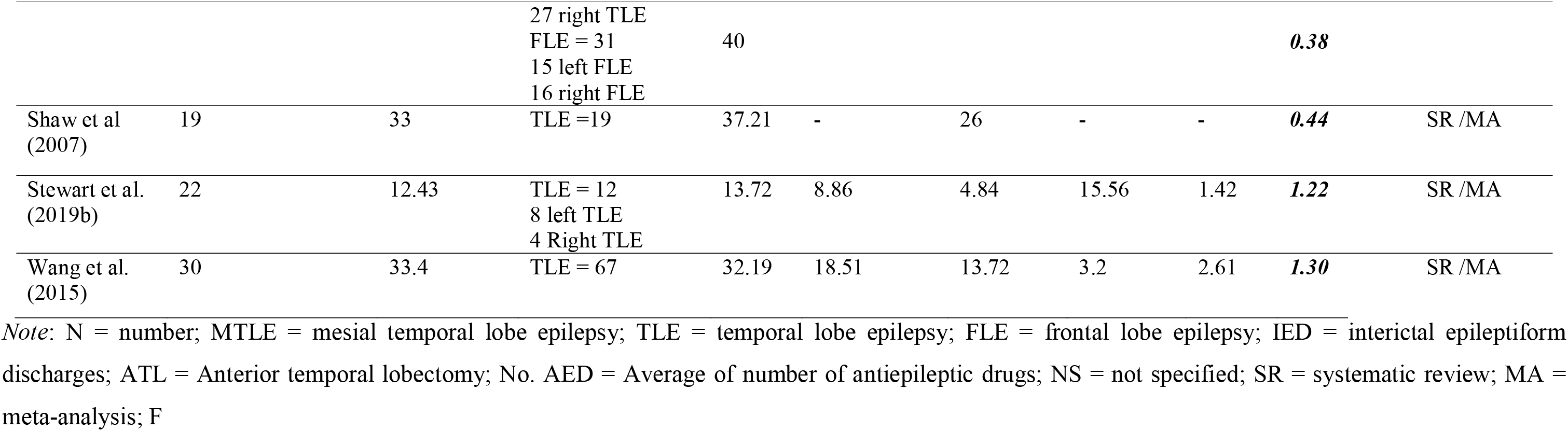
Study characteristics and effect sizes (Hedge’s g) reported individually for studies included in the meta-analysis pertaining theory of mind (ToM)

*Empathy*: We included studies that looked at the generalized construct as well as the cognitive and affective sub-components that assesses how individuals perceive others’ emotions and perspective and how they share emotional states with others, respectively. All eligible studies evaluating empathy and their effect sizes are reported in Table 4.

**Table 4.** Study characteristics and effect sizes (Hedge’s g) reported individually for studies included in the meta-analysis pertaining to empathy and social behavior.

| Study | Non-clinical group |  | Epilepsy group |  |  |  |  |  | Hedge's g | Included in the SR / MA |
| --- | --- | --- | --- | --- | --- | --- | --- | --- | --- | --- |
|  | N | Age | N | Age | Age of onset (years) | Duration (years) | Seizures p/month | No. AED/ |  |  |
| Empathy |  |  |  |  |  |  |  |  |  |  |
| Broicher et al. (2012b) | 29 | 33.69 | MTLE = 28 | 34.43 | 20.21 | 14.25 | - | - | 0.17 | SR /MA |
| Gul & Ahmad et al. (2017) | 60 | 28.83 | FLE = 60 | 28.7 | 13.23 | - | - | - | - | SR |
| Hennion et al. (2015a) | 50 | 42.81 | TLE = 50<br>27 left TLE<br>23 Right TLE | 42.50 | 21.06 | 21.34 | 12.32 | 2.14 | 0.41 | SR /MA |
| Hu et al. (2016) | 60 | 17.18 | Frontal IED = 10<br>43 extrafrontal | 18.87 | 15.14 | 3.74 | - | - | 0.96 | SR /MA |
| Realmuto et al. (2015) | 21 | 31.95 | TLE = 21 | 37 | 24.3 | 12.9 | - | 1.3 | 0.69 | SR /MA |
| Toller et al. (2015a) | 30 | 38.2 | MTLE = 34<br>16 left TLE<br>18 right TLE<br>16 extra-MTLE | 38.3 | 11.5 | 21.5 | - | - | 0.83 | SR /MA |
| Toller et al. (2015b) | 33 | 39.1 | MTLE = 22<br>9 left TLE<br>13 right TLE | 36.65 | 16.3 | 20.75 | - | - | 0.83 | SR /MA |
| Social behavior |  |  |  |  |  |  |  |  |  |  |
| Gois et al. (2011) | 38 | 28.61 | TLE = 35<br>16 right TLE<br>19 right TLE | 39.82 | 13.89 | 25.91 | 54.3 more<br>45.7 less | 7 on mono<br>28 on | -0.90 | SR /MA |

Social cognition and Epilepsy
|  |  |  |  |  |  |  | than 1 | poly |  |  |
| --- | --- | --- | --- | --- | --- | --- | --- | --- | --- | --- |
| Gascoigne et al. (2019) | 58 | - | TLE = 23<br>6 right TLE<br>15 left TLE | 11.7 | 5.71 | 5.36 | - | 1.3 | **-1.39** | SR /MA |
| Lunn et al. (2015) | 62 | 10.5 | TLE = 6<br>FLE = 5 | 11.5 | 6.5 | 4 | 2 | 18 on<br>mono 5<br>on poly | **1.28** | MA |
| Stewart et al. (2019a) | 22 | 12.43 | TLE = 12<br>8 left TLE<br>4 Right TLE | 13.72 | 8.86 | 4.84 | 15.56 | 1.42 | **0.63** | SR /MA |
| Stewart et al. (2019b) | 22 | 12.43 | TLE = 12<br>8 left TLE<br>4 Right TLE | 13.72 | 8.86 | 4.84 | 15.56 | 1.42 | **0.63** | SR /MA |

*Note:* N = number; MTLE = mesial temporal lobe epilepsy; TLE = temporal lobe epilepsy; FLE = frontal lobe epilepsy; IED = interictal epileptiform discharges; No. AED = Average of number of antiepileptic drugs; NS = not specified; SR = systematic review; MA = meta-analysis.

*Social behavior*: Given that social behavior has been operationalized differently in the literature, we only included studies that employed one or more widely used measures considered relevant to this construct such as Social Responsiveness Scale, Child Behavior Checklist (CBCL) and Social Adjustment Scale. All eligible studies evaluating social behavior and their effect sizes are reported in Table 4.

### Data extraction and data entry

We extracted the available participant characteristics (e.g., mean age, standard deviation and number of participants) in each clinical and non-clinical control group for each study. For clinical groups, we recorded the mean age of onset, duration of epilepsy, and the number of seizures per month, where available. For each social cognition domain, the mean and standard deviation of items reported (accuracy or errors) for each task was extracted for each clinical and non-clinical control group. Where the mean of errors was reported (Amlerova et al., 2014; Cohn et al., 2015; Laurent et al., 2014; Schacher et al., 2006), the effect size calculation was reversed to account for this difference in the direction of effect. Alternatively, for studies where raw values were not available, other statistics such as events rates (Gomez-ibanez et al., 2014), *p* values and sample size were used to calculate effect sizes (Amlerova et al., 2014; Batut et al., 2018; Meletti et al., 2003b; Schacher et al., 2006). We contacted authors to request subgroup raw values when focal epilepsy groups or laterality data were combined. Data were made available in several instances (Cohn et al., 2015; Laurent et al., 2014; Tanaka et al., 2013; Toller et al., 2015a & 2015b; Stewart et al., 2019a & 2019b; Bujarski et al., 2016; Morou et al., 2018; Lunn et al., 2015). Studies that only included post-operative patients were excluded from the estimation of effect sizes. Data published more than 10 years ago were assumed to be unavailable.

For studies with more than one outcome measure in each social-cognitive domain, effect sizes were pooled to calculate an overall effect for each construct. If studies reported both pre- and post-operative data, only pre-operative data were included. For studies in which different intensities of emotional expression were used to evaluate emotion recognition, we included the data for the 100% intensity of facial expression in the meta-analyses and the remaining intensity data were narratively synthesised (Sedda et al., 2013). For studies in which different task instructions were used, the index reported by the authors was included in the analysis (Shaw et al., 2007). Due to their limited number, studies employing widely used measures of social behavior and empathy are discussed as part of the systematic review but results from meta-analyses are provisional and require replication in future. First author extracted the data and second author, CA, checked the accuracy of extracted data independently.

Tables 2-4 present a study-by-study breakdown of effect sizes for each domain of social cognition in patients compared to non-clinical controls.

### Quality rating

The Downs and Black Checklist (1998) was used to evaluate the methodological quality of included studies in the meta-analysis. The checklist includes measures for the psychometric properties of the papers for randomized and non-randomized studies. The checklist shows good test-retest reliability (*r*= .88), inter-rater reliability (*r*= .75), and internal consistency (Kruder-Richardson formula 20= .89; Downs and Black, 1998). The adapted 17-item version of this checklist was used; items related to interventional trials were omitted as studies of this nature were not included for review. This checklist evaluates the quality of reporting (items 1-8), external validity (item 9), internal validity (statistical and methodological bias, items 10-12; selection bias, items 13-16), and power (item 17). All items are scored from 0 (no, or unable to be determined) to 1 (yes), except for item 4, which is scored 0 (no, unable to be determined), 1 (partially), or 2 (yes). Items 7 and 16 were only scored for studies with a longitudinal design. Therefore, scores for cross-sectional studies, ranged from 0-16, while longitudinal studies scores ranged from 0-18. Studies were categorized into three categories with high (0-5 points for cross-sectional and longitudinal studies), average (6-10 points for cross-sectional and 6-11 points for longitudinal), and low (11-16 points for cross-sectional and 12-18 points for longitudinal studies) risk of bias. A similar procedure to that reported by Edwards et al., 2017; Stewart et al., 2016 was used for quality rating using this checklist. Two authors (MZ & CA) independently reviewed and scored all papers and discrepancies were resolved through discussion.

### Methods of Review

Figure 1 displays the flow diagram describing the process of study selection for the review. The initial search retrieved 6181 articles. After duplicates were removed, 3847 titles and abstracts were screened for the relevance and eligibility of the papers, 108 full texts were assessed for the final eligibility independently by two authors (90% agreement). Forty-four eligible papers remained for the systematic review, 40 of which were included for meta-analysis.

**Figure 1.**
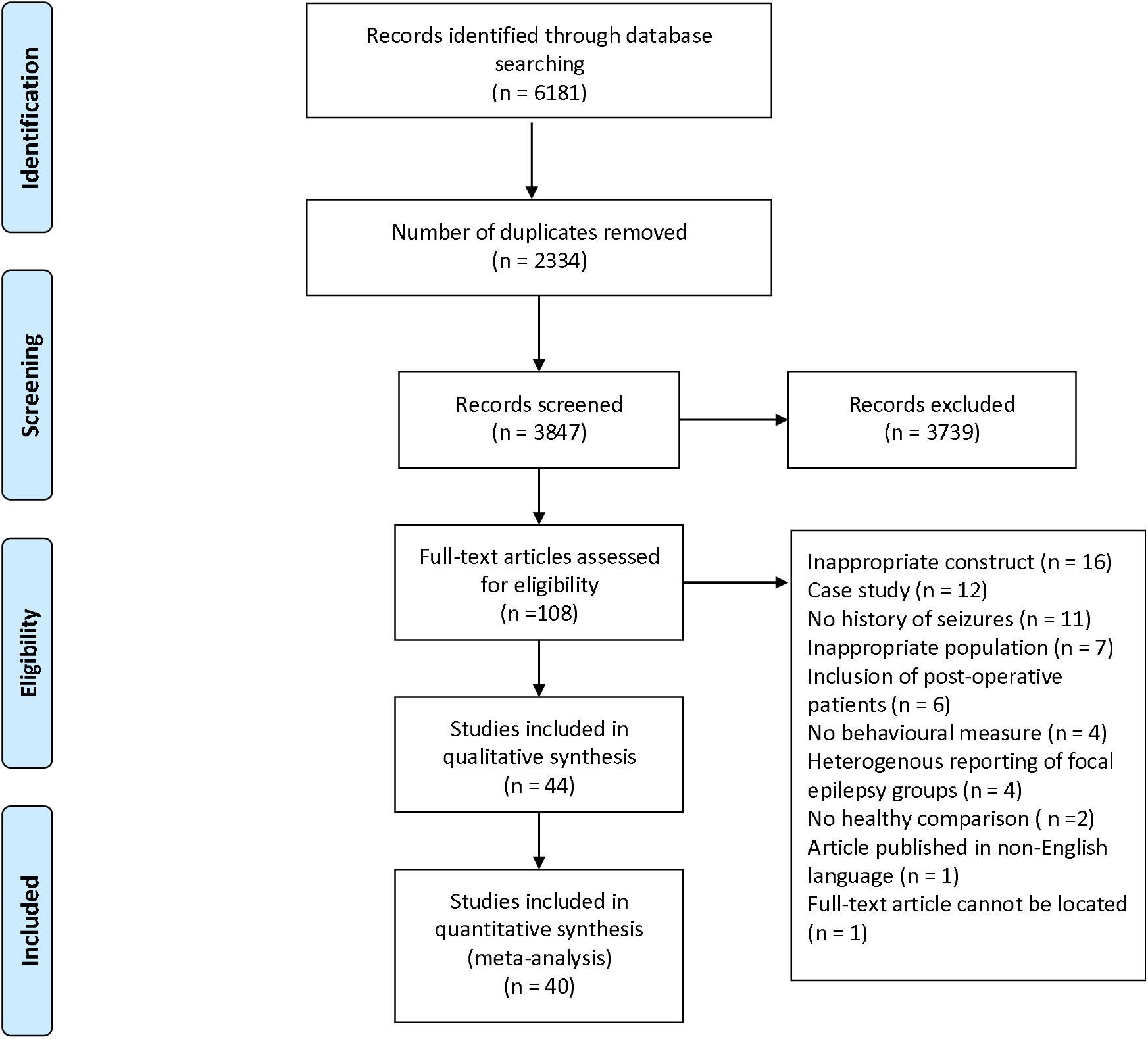
Flow diagram of identification and selection procedure of studies based on PRISMA guideline

### Patient Demographics

Overall studies included 2830 non-clinical control participants and 1595 patients with epilepsy, 113 patients with FLE and 1482 patients with TLE. The age ranged from 10 to 48 years in the non-clinical group and 12 to 52 years in the patient group. For the patient group, the mean age-of-onset of epilepsy was 13 years for the emotion recognition studies, 13 years for theory of mind studies, 17 years for the empathy studies, and 20 years for social behavior studies. The average duration of epilepsy was 12 years for emotion recognition studies, 11 years for theory of mind studies, 16 years for empathy studies, 10 years for social behavior studies.

### Statistical analysis

Data were analyzed using the Comprehensive Meta Analyses Program, Version 3 (Borenstein et al., 2013). Effect sizes were calculated as Hedges’ *g.* We interpreted the importance and strength of our effect in line with the Cohen’s *d* guidelines, with 0.2, 0.5, and 0.8 corresponding to small, medium, and large effects, respectively (Cohen, 1988). A negative effect indicates that the epilepsy group performed more poorly than non-clinical controls and positive effect indicates the reverse. Effect size by task type was also compared using the *Q*-test, indicating the degree of variability within each effect in question where the larger values indicate larger between-groups variability. In other words, the *Q* value larger than the number of pooled effects (k-1 degree of freedom) indicates that differences between groups were significant within each task domain.

The analyses examined the overall and specific effects for each social cognitive construct: (1) overall difference in social cognition performance (emotion recognition, theory of mind and empathy) between TLE and FLE compared to non-clinical controls, and between TLE subgroups (side of seizure focus - left versus right); (2) performance for specific emotions (anger, sadness, happiness, disgust, and fear) between TLE and FLE compared to non-clinical controls, and between TLE subgroups (side of seizure focus - left versus right); (3) performance for sub-components of theory of mind (e.g., Faux Pas and False Belief tasks) between TLE and FLE compared to non-clinical controls, and between TLE subgroups (side of seizure focus - left versus right). Due to known variation, random-effect models were used for all analyses to ensure that we captured the heterogeneous effects present in clinical populations. Where permitted, the effects of laterality of the seizure focus on task performance (e.g., visual vs. auditory emotion recognition) were assessed in each social cognitive domain.

Initial analyses included calculation of the weighted mean effect size for each domain of social cognition (e.g., a single effect size for each dependent measure for each independent study). Subsequent analyses focused on left and right-side seizure focus vs. non-clinical control participants in each domain of social cognitive function. Finally, exploratory analyses were conducted on specific theory of mind tasks for TLE vs. non-clinical controls and specific emotions that are reported at the end.

### Results of systematic review & meta-analyses

#### Emotion Recognition

##### Temporal Lobe Epilepsy (TLE)

###### Facial emotion recognition

Twenty-three studies examined facial emotion recognition in TLE patients, including the use of static and dynamic stimuli as well as stimuli displaying facial expressions at varying intensities (Table 1). Among these studies, 17 of 18 reported impaired emotion recognition using a composite score. Of the individual emotions, deficits were most commonly reported in fear recognition (11 of 18 studies), followed by disgust (10 of 17 studies) and then sadness (6 of 19 studies) in all types of stimuli (e.g., dynamic or static). Two of 15 studies found deficits in recognising angry facial expressions, whereas only one of 18 studies found deficits in recognising happy facial expressions.

###### Lateralisation

Eleven of the above studies reported effects of laterality of the epileptic focus, compared to non-clinical controls (seven included right TLE (RTLE) and six left TLE (LTLE)). For overall emotional recognition ability, four of five studies found impairments in RTLE patients and three of five studies found impairments in LTLE patients compared to non-clinical controls. In terms of recognising specific emotions, three of seven studies examining fear demonstrated deficits in RTLE and one of seven studies demonstrated impairments in LTLE. Two of six studies showed that RTLE patients were impaired in their recognition of sadness (Melleti et al., 2003a; Melleti et al., 2003b) whereas LTLE patients exhibited intact recognition of sadness compared to non-clinical controls (Seven of seven studies). One of six studies measuring happiness found that RTLE patients were impaired in their recognition of happiness (Cohn et al., 2015); however, no deficits emerged for LTLE patients among all seven studies (See Table 6). Seven studies compared the performance between right-side epilepsy patients and controls in recognizing disgust with three of these studies reporting deficits in RTLE patients (Golobouff et al., 2008; Melletti et al., 2003a; Melletti et al., 2003b) whereas none of six reported deficits in LTLE patients. None of the five studies examining anger found impairments in either RTLE or LTLE patients.

Thirteen studies made direct comparisons between LTLE and RTLE in facial emotion recognition. A minority of the studies that reported the composite score found lateralised differences in total emotion recognition (three of 11), with RTLE patients performing worse than LTLE patients (Meletti et al., 2003b; Meletti et al. 2009; Sedda et al., 2013). Meletti et al. (2009) reported that bilateral TLE (BTLE) patients performed worse than RTLE patients, and RTLE performed worse than LTLE patients. Additionally, Sedda et al., (2013) found that RTLE patients were more impaired at recognising emotions at 75% intensity than LTLE patients; however, these differences were non-significant when expressions were presented at 100%.

Eight studies examined fear recognition in patients with RTLE and LTLE. Five of these studies found differences according to seizure lateralisation. Specifically, four studies found that RTLE patients were more impaired in fear recognition than LTLE patients. Meletti et al. (2009) showed that fear recognition was comparable in BTLE and RTLE patients, with both groups performing worse than LTLE patients. In contrast, one study reported that LTLE patients performed worse than RTLE patients in fear recognition (Golobouff et al., 2008). Three studies found a non-significant difference (Reynders et al., 2005; Stewart et al., et al., 2019; Shaw et al., 2007).

Finally, six studies inspected the differences in recognising other emotions (i.e., happiness, anger, sadness and disgust), in addition to fear, according to seizure lateralisation. One study reported significant differences between RTLE and LTLE patients in the recognition of anger and sadness (Meletti et al., 2009). Meletti et al. (2009) found that BTLE and RTLE patients performed equally in recognizing anger and sadness but were worse than LTLE patients. No differences were found according to seizure laterality for happiness or disgust.

Results from the meta-analysis on all included emotion recognition studies in TLE produced a Hedge’s *g* of 0.69 and a 95% confidence interval (CI) for the effect of between 0.35 to 1.04; Z score = 3.93, *p* < 0.001; Table 5). As this range does not include zero, the effects are considered significantly different from zero. Therefore, we can reject the null hypothesis and conclude that TLE patients significantly and substantially performed worse than non-clinical groups on emotion recognition tasks.

Additionally, TLE patients with left versus right-sided seizure lateralisation were compared to a non-clinical population. Our meta-analysis suggested that the small mean effect for LTLE (Hedge’s *g* = 0.35; 95% CI = −0.01-0.68; Z score = 2.06, *p* =0.039) and RTLE (Hedge’s *g* = 0.42; 95% CI = −0.04-0.80; Z score = 2.21, *p* =0.027; Table 5) compared to the non-clinical group. Given that the CIs cross zero, the effects are not significant, suggesting that there is no statistical difference in emotion recognition comparing the LTLE and RTLE to non-clinical group.

##### Frontal Lobe Epilepsy (FLE)

Overall, three studies have looked at facial emotion recognition in FLE (Farrant et al., 2005; Golouboff et al., 2008; Hu et al., 2016). Two studies evaluated overall emotional recognition ability, one study revealing impairments in FLE (Farrant et al., 2005) while the other did not (Golouboff et al., 2008; Hu et al., 2016). In terms of specific emotions, a greatest number of impairments was found in the recognition of anger, fear and sadness (two of three: Farrant et al., 2005; Hu et al., 2016); However, Golouboff et al. (2008) did not find impairments in these emotions. Only one of three studies found deficits in the recognition of happiness (Farrant et al., 2005) and disgust (Hu et al., 2016). Differences between sides were not reported in any of the studies.

At meta-analytic review, a large effect size was achieved for emotion recognition in FLE (Hedge’s *g* = 1.18; 95% CI = 0.56-1.80). The Z-value for testing the null hypotheses was 3.71 for emotion recognition (*p*-value < 0.001; Table 5), suggesting that performance was poorer in FLE than in non-clinical controls, however, the results need to be considered cautiously due to small number of studies and participants in this category.

**Table 5.**
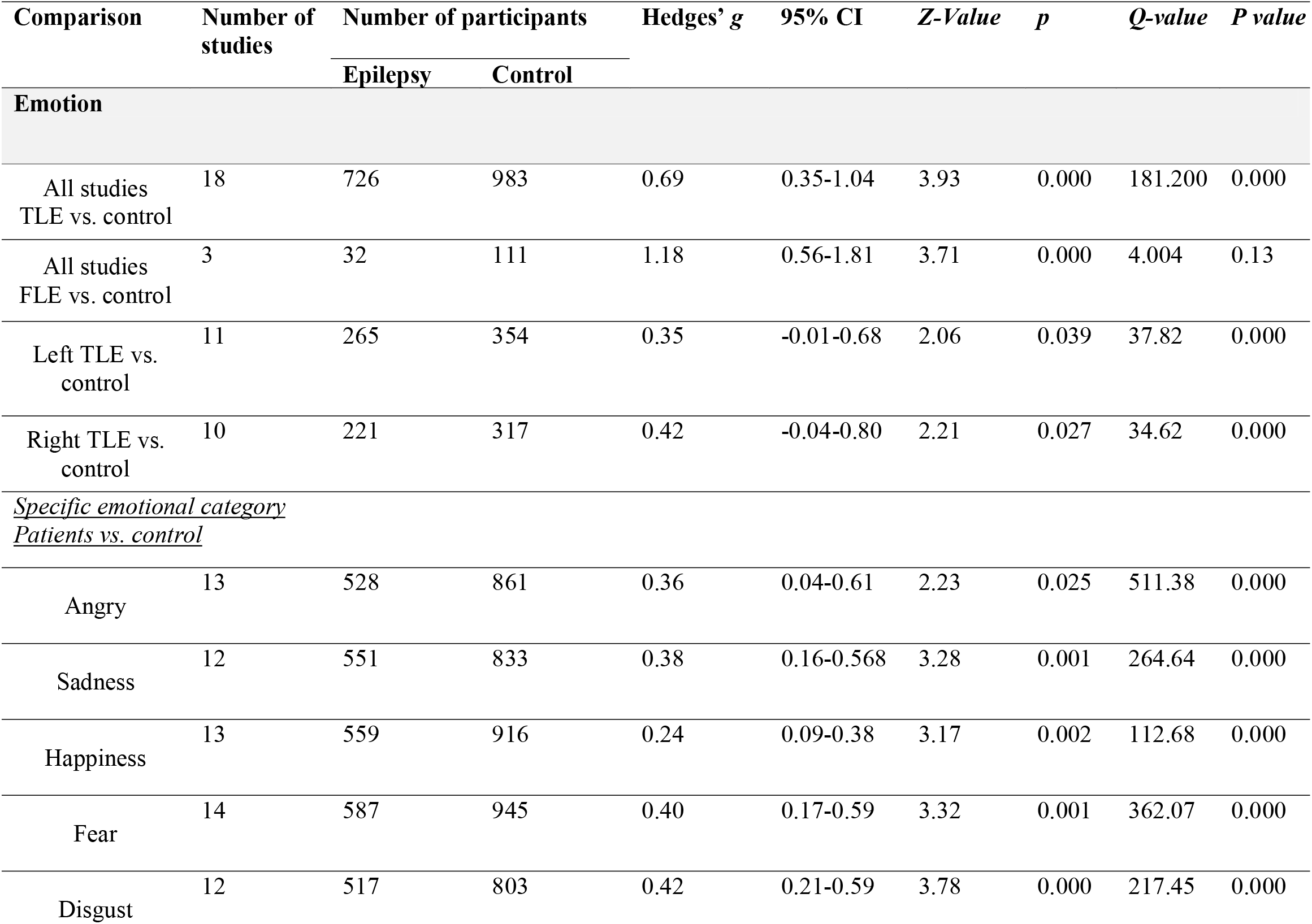
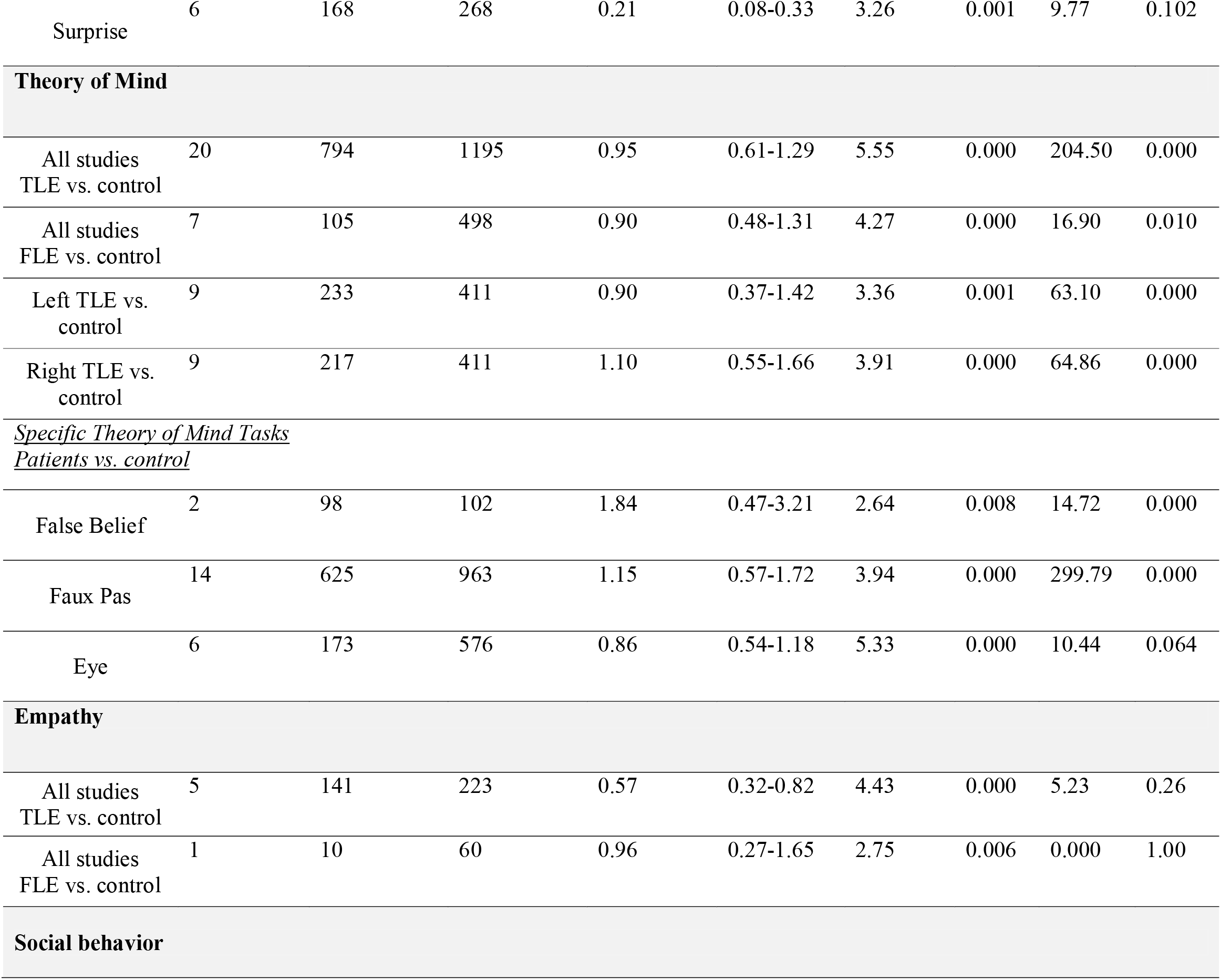
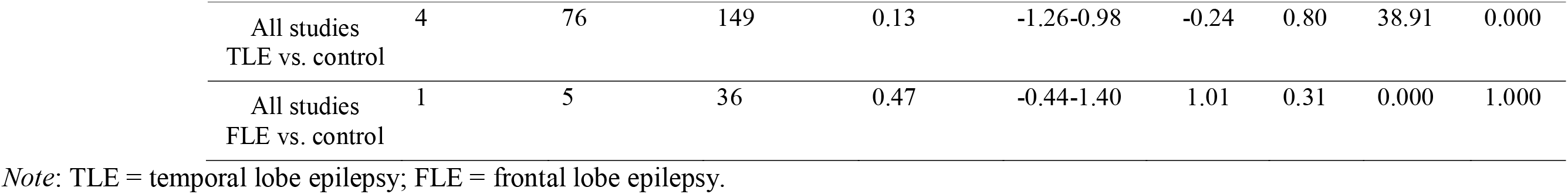
Hedges’ g for social cognitive measures, comparing control with temporal and frontal lobe epilepsy. Data are presented separately according to each social cognitive domain and for each patients’ subgroups as well as subtypes within each social cognitive domain.

| Comparison | Number of studies | Number of participants |  | Hedges' g | 95% CI | Z-Value | p | Q-value | P value |
| --- | --- | --- | --- | --- | --- | --- | --- | --- | --- |
|  |  | Epilepsy | Control |  |  |  |  |  |  |
| Emotion |  |  |  |  |  |  |  |  |  |
| All studies<br>TLE vs. control | 18 | 726 | 983 | 0.69 | 0.35-1.04 | 3.93 | 0.000 | 181.200 | 0.000 |
| All studies<br>FLE vs. control | 3 | 32 | 111 | 1.18 | 0.56-1.81 | 3.71 | 0.000 | 4.004 | 0.13 |
| Left TLE vs.<br>control | 11 | 265 | 354 | 0.35 | -0.01-0.68 | 2.06 | 0.039 | 37.82 | 0.000 |
| Right TLE vs.<br>control | 10 | 221 | 317 | 0.42 | -0.04-0.80 | 2.21 | 0.027 | 34.62 | 0.000 |
| <u>Specific emotional category</u><br><u>Patients vs. control</u> |  |  |  |  |  |  |  |  |  |
| Angry | 13 | 528 | 861 | 0.36 | 0.04-0.61 | 2.23 | 0.025 | 511.38 | 0.000 |
| Sadness | 12 | 551 | 833 | 0.38 | 0.16-0.568 | 3.28 | 0.001 | 264.64 | 0.000 |
| Happiness | 13 | 559 | 916 | 0.24 | 0.09-0.38 | 3.17 | 0.002 | 112.68 | 0.000 |
| Fear | 14 | 587 | 945 | 0.40 | 0.17-0.59 | 3.32 | 0.001 | 362.07 | 0.000 |
| Disgust | 12 | 517 | 803 | 0.42 | 0.21-0.59 | 3.78 | 0.000 | 217.45 | 0.000 |
| Surprise | 6 | 168 | 268 | 0.21 | 0.08-0.33 | 3.26 | 0.001 | 9.77 | 0.102 |
| **Theory of Mind** |  |  |  |  |  |  |  |  |  |
| All studies<br>TLE vs. control | 20 | 794 | 1195 | 0.95 | 0.61-1.29 | 5.55 | 0.000 | 204.50 | 0.000 |
| All studies<br>FLE vs. control | 7 | 105 | 498 | 0.90 | 0.48-1.31 | 4.27 | 0.000 | 16.90 | 0.010 |
| Left TLE vs.<br>control | 9 | 233 | 411 | 0.90 | 0.37-1.42 | 3.36 | 0.001 | 63.10 | 0.000 |
| Right TLE vs.<br>control | 9 | 217 | 411 | 1.10 | 0.55-1.66 | 3.91 | 0.000 | 64.86 | 0.000 |
| <u>Specific Theory of Mind Tasks</u><br><u>Patients vs. control</u> |  |  |  |  |  |  |  |  |  |
| False Belief | 2 | 98 | 102 | 1.84 | 0.47-3.21 | 2.64 | 0.008 | 14.72 | 0.000 |
| Faux Pas | 14 | 625 | 963 | 1.15 | 0.57-1.72 | 3.94 | 0.000 | 299.79 | 0.000 |
| Eye | 6 | 173 | 576 | 0.86 | 0.54-1.18 | 5.33 | 0.000 | 10.44 | 0.064 |
| **Empathy** |  |  |  |  |  |  |  |  |  |
| All studies<br>TLE vs. control | 5 | 141 | 223 | 0.57 | 0.32-0.82 | 4.43 | 0.000 | 5.23 | 0.26 |
| All studies<br>FLE vs. control | 1 | 10 | 60 | 0.96 | 0.27-1.65 | 2.75 | 0.006 | 0.000 | 1.00 |
| **Social behavior** |  |  |  |  |  |  |  |  |  |
| All studies<br>TLE vs. control | 4 | 76 | 149 | 0.13 | -1.26-0.98 | -0.24 | 0.80 | 38.91 | 0.000 |
| All studies<br>FLE vs. control | 1 | 5 | 36 | 0.47 | -0.44-1.40 | 1.01 | 0.31 | 0.000 | 1.000 |

*Note:* TLE = temporal lobe epilepsy; FLE = frontal lobe epilepsy.

#### Theory of Mind

##### Temporal Lobe Epilepsy

Seventeen studies included at least one measure of Theory of Mind with the Faux Pas test being the most used task (Table 3). None of the 17 studies that examined Theory of Mind in focal epilepsy provided composite scores; thus, the results of each task will be examined separately. For the Faux Pas task, 12 of 13 studies reported lower performance in TLE patients than in non-clinical controls. Regarding seizure lateralisation, four of four studies found that both RTLE and LTLE patients were impaired compared with non-clinical controls, and one study also showed similar performance in BTLE compared to RTLE and LTLE patients (Li et al., 2013). Furthermore, one of four studies found that RTLE patients were more impaired than LTLE patients (Broichera et al., 2012a), and the remaining studies reported a non-significant difference according to seizure lateralisation (Broicher et al., 2012b; Hennion et al., 2015b; Shaw et al., 2007; Stewart et al., 2019; see supplementary table 2 for breakdown of studies).

For the False Beliefs task, two of three studies found that patients with TLE scored lower than non-clinical controls (Wang et al., 2015; Li et al., 2013). One study comparing epileptic seizure lateralisation to non-clinical controls reported that RTLE and BTLE patients were impaired but LTLE was not impaired (Li et al., 2013). For the Reading the Mind in the Eye test, one of two studies reported that mesial TLE patients were more impaired than non-clinical controls (Okruzek et al., 2017), and no study identified significant differences according to seizure lateralisation (Broichera et al., 2012b).

Of the studies that have used the Strange Stories test, four studies found that TLE patients were more impaired than controls (Li et al., 2013, Shaw et al., 2004, Stewart et al., 2019; Wang et al., 2015) while one study did not find any difference (Shaw et al., 2007). In the only study comparing TLE subgroups according to seizure lateralisation with non-clinical controls, deficits were reported amongst RTLE patients, but not LTLE or BTLE patients (Li et al., 2013). However, no significant differences emerged between patients with epilepsy grouped by seizure lateralisation and controls on this task in two studies (Shaw et al., 2007; Stewart et al., 2019).

All three studies that used the moving triangles task reported that mesial TLE patients performed worse than non-clinical controls (Bala et al., 2018; Broichera et al., 2012b; Hennion et al., 2016). Only one study reported that both right and left mesial TLE patients performed worse than non-clinical controls (Hennion et al., 2016); however, when comparing patients according to seizure lateralisation, left and right-TLE patients performed comparably (Broichera et al., 2012b; Hennion et al., 2016). Additionally, Hennion et al. (2016) reported that brain activation in a ToM-interaction condition relative to non-theory of mind interaction condition showed greater activation of inferior and middle occipital gyrus in right mesial TLE than in non-clinical controls. Non-clinical control participants, however, activated inferior and middle occipital gyrus, left temporoparietal junction, fusiform gyri, and superior temporal sulcus more than patients.

Results from meta-analysis suggested that the effect for theory of mind was large (Hedge’s *g* = 0.95; 95% CI = 0.61-1.29; Z score = 5.55, *p* < 0.001; Table 5). As the confidence interval does not include zero, the effects were significantly different from zero; hence, we can reject null hypotheses and conclude that on average, TLE patients performed significantly, and substantially, worse than non-clinical groups on theory of mind. Additionally, our results indicated large effect sizes for both right and left lateralized TLE. However, the magnitude of this impairment was larger for patients with RTLE (Hedge’s *g* = 1.10; 95% CI = 0.55-1.66; Z score = 3.91, *p* < 0.001) compared to LTLE (Hedge’s *g* = 0.90; 95% CI = 0.37-1.42; Z score = 3.36, *p* =0.001). The difference between the two means yields a Q-value of 64.86 for RTLE (*p* < 0. 001) and 63.10 for LTLE (*p* < 0. 001) (Table 5). We reject the null hypotheses that the means of the two groups are identical and conclude that patients had more severe theory of mind difficulties than non-clinical controls and also that RTLE patients had more deficits than LTLE patients. Table 6 summarizes the results for comparison between LTLE and RTLE in emotion recognition and theory of mind constructs.

**Table 6.** Comparison of temporal lobe epileptic lesion sides to non-clinical controls and to each other in emotion recognition and theory of mind ability.

| Author | LTLE vs HC | RTLE vs HC | LTLE VS RTLE |
| --- | --- | --- | --- |
| **Emotion Recognition** |  |  |  |
| ***Visual/Facial stimuli*** |  |  |  |
| Amlerova | □□ = T | □□ = T | □□ = T |
| Batut | □□ = T, F | □□ = T, F | □□ = T |
| Bonora | - | - | □□ = T |
| Broicher | - | - | □□ = F (RTLE < LTLE)<br>□□ = T, H, A, S, D |
| Cohn | □□ = T<br>□□ = H, S, D | □□ = T, H<br>□□ = S, D | - |
| Fowler | □□ = H, A, F, S, D | □□ = H, A, F, S, D | - |
| Golouboff | □□ = T, F<br>□□ = H, A, S, D | □□ = D<br>□□ = T, H, A, F, S | □□ = F (LTLE < RTLE)<br>□□ = T, H, A, S, D |
| Hennion (2016) | - | - | □□ = T |
| Hlobil | - | □□ = F | □□ = F (RMTLE < LMTLE) |
| Laurent | - | - | □□ = T |
| Meletti (2003a) | - | □□ = S, D | - |
| Meletti (2003b) | □□ = H, A, F, S, D | □□ = F, S, D<br>□□ = H, A | □□ = T, F (RTLE < LTLE) |
| Meletti (2009) | - | - | □□ = T (BTLE < RTLE < LTLE); S, F, A<br>(BLTE = RTLE < LTLE).<br>□□ = H, D |
| Reynders | - | - | □□ = F |
| Sedda | **35%, 50%, 75%, 100% intensity:**<br>□□ = T, H, A, F, S, D | **35%, 50%, 75% intensity:**<br>□□ = T<br>□□ = H, A, F, S, D<br>**100% intensity**<br>□□ = T, H, A, F, S, D | **35%, 50% intensity:**<br>□□ = T, H, A, F, S, D<br>**75% intensity:**<br>□□ = T (RTLE < LTLE)<br>□□ = H, A, F, S, D |
| Shaw | □□ = H, A, F, S, D | □□ = H, A, F, S, D | □□ = H, A, F, S, D |
| Stewart | - | - | □□ = T, H, A, F, S, D |
| Szaflarski | □□ = H, F, S | - | - |
| ***Auditory*** |  |  |  |
| Bonora | - | - | □□ = T |
| Fowler | □□ = T, H, A, F, S, D | □□ = T, H, A, F, S, D | - |
| Hennion | - | - | □□ = T |
| Laurent | - | - | □□ = T |
| Author | LTLE vs HC | RTLE vs HC | LTLE VS RTLE |

Social cognition and Epilepsy
| **Theory of Mind** |  |  |  |
| --- | --- | --- | --- |
| Amlerova | □□ = FP | □□ = FP | - |
| Broicher 2012a | - | - | □□ = FP (RMTLE < LMTLE) |
| Broicher 2012b | - | - | □□ = FP, RMET, MS, |
| Cohn | - | - | □□ = TASIT deceitful exchanges (RTLE < LTLE) |
| Giovagnoli 2011 | □□ = FP | □□ = FP | - |
| Giovagnoli 2016 | □□ = FP | □□ = FP | - |
| Hennion 2016 | □□ = MT | □□ = MT | □□ = MT |
| Li | □□ = FP, CTN implicit form<br>□□ = FB, SS, CTN explicit form | □□ = FP, FB, SS, CTN implicit & explicit form | - |
| Shaw 2004 | - | - | □□ = Cumulative ToM score (i.e., FP, FB, SS, MIT, CBET) |
| Shaw 2007 | - | - | □□ = FP, SS |
| Stewart (2019) | - | - | □□ = FP, SS |

Studies are included if they compared the different lesion sides to non-clinical controls and to each other. LTLE = Left temporal lobe epilepsy. RTLE = Right temporal lobe epilepsy. BTLE = Bilateral temporal lobe epilepsy. T = total score; H = happy; A = angry; F = fearful; S = sadness; D = disgust; FP = faux pas task; FB = false belief task; RMET = reading mind in the eyes task; MS = moving shapes task; SS = strange stories task; CTN = cartoon vignettes task; MT = moving triangle; % = Intensity of the facial expression presented in Sedda et al. (2013); □□ = significant difference ( $p < .05$ ); □□ = non-significant difference.

##### Frontal Lobe Epilepsy

Overall, studies that have evaluated theory of mind in FLE patients reported lower performance on the Faux Pas test (two of three studies) the RMET (two of two studies), and cartoon vignettes test (only one study) compared to non-clinical controls (Table 3). One study that used the Yoni task found that FLE patients performed worse than controls on the sub-measures of second-order ToM, “fortune of others”, envy, gloating, and identification (Hu et al., 2016). However, FLE patients were not impaired relative to controls on the Strange Stories task (Farrant et al., 2005). At meta-analytic review, a large effect size was achieved for theory of mind (Hedge’s *g* = 0.90; 95% CI = 0.48-1.31). The Z-value for testing the null hypotheses was 4.27 (*p*-value < 0.001), suggesting that patients with FLE performed more poorly than non-clinical controls in ToM. Due to small number of studies and participants, these results need to be interpreted cautiously.

#### Empathy

##### Temporal Lobe Epilepsy

Four studies measured empathy in TLE patients (Broicher et al., 2012b; Hennion et al., 2015b; Toller et al., 2015a; Toller et al., 2015b), and each study reported a different combination of the total score and subscale scores (Table 4). Two studies looking at the total empathy score found that TLE patients were unimpaired compared to non-clinical controls (Hennion et al., 2015b; Broicher et al., 2012b). Regarding the cognitive component of empathy, one study revealed deficits in TLE patients (Hennion et al., 2015b) while another did not (Broicher 2012b). For the perspective-taking subscale, deficits were reported in RTLE patients compared with non-clinical controls (Toller et al., 2015b). However, no deficits were found in the fantasy subscale (Broicher et al., 2015b). No differences between TLE groups according to seizure lateralisation were found on the total score, cognitive component score, or fantasy subscore (Broicher et al. 2012b), or perspective-taking subscore (Toller et al., 2015b).

In studies examining various aspects of the affective component, no impairments were found in the total affective score (Broicher et al., 2012b; Hennion et al., 2015b). Two of three studies reporting empathic concern found deficits in RTLE but not LTLE patients (Toller et al., 2015a; Toller et al., 2015b). For the personal distress scale, one of two studies showed that RTLE and LTLE patients had higher scores than non-clinical controls (Toller et al., 2015a). Three studies compared RTLE and LTLE patients. RTLE patients were impaired in empathic concern relative to LTLE patients in two of the three studies (Toller et al., 2015a; Toller et al., 2015b), whereas no differences between sides were reported in personal distress (two of two studies) (Broicher et al., 2012b; Toller et al., 2015a) or in the overall affective component (one of one study) (Broicher et al., 2012b). Results from meta-analysis revealed that for the empathy domain the effect size was moderate with a Hedge’s g of 0.57 (95% CI = 0.32-0.82; Z score = 4.43, *p* < 0.001; Table 5). As the confidence interval does not include zero, the effects were significantly different from zero, so, we can reject the null hypotheses and conclude that on average, TLE patients performed significantly, and substantially, worse than non-clinical groups on empathy.

##### Frontal Lobe Epilepsy

Two studies measured empathy in FLE patients (Gul & Ahmad, 2017; Hu et al., 2016). One study reported the total score from the IRI, which showed that FLE patients were impaired in empathy relative to non-clinical controls (Hu et al., 2016). Both studies showed impairments in cognitive empathy but no impairments in affective empathy. Of the two studies, only Hu et al. (2016) reported results of the four cognitive subscales (i.e., perspective taking, fantasy, empathic concern, and personal distress). Deficits were found on the perspective taking subscale but not the fantasy subscale, and no impairments were found on either empathic concern or personal distress. Differences between FLE patients grouped according to seizure lateralisation were not reported in any study (Table 4).

Due to the low number of studies in the analysis, statistical measures need to be interpreted cautiously. Overall, we found high Hedge’s g of 0.96 (95% CI = 0.27-1.65; Z score = 2.75, *p* < 0.001; Table 5) suggesting that FLE patients had lower empathic response than non-clinical population.

#### Social behavior

##### Temporal Lobe Epilepsy

Studies included in this systematic review employed only three scales: Social Responsiveness Scale, Child Behavior Checklist (CBCL) and Social Adjustment Scale. Four studies compared TLE patients with non-clinical controls on different aspects of social behavior (Gois, et al., 2011; Gascoigne, et al., 2019; Stewart et al., 2019a; Stewart, et al., 2019b). One study investigated social adjustment in adults with TLE compared to non-clinical controls (Gois, et al., 2011) and found impairments in overall social adjustment as well as subdomains pertaining to work and leisure. However, the remaining subdomains (i.e., family relationship, marital relationship, relationship with children, domestic life, and financial situation) were unaffected in patients with TLE (Table 4).

The remaining three studies investigated social behavior in children and adolescents. Two studies assessed participants on the Social Competence Scale from the Child Behaviour Checklist (CBCL), in which one reported impairment (Stewart, et al., 2019b) whereas the other did not (Stewart, et al., 2019a). Another study assessed and reported significantly elevated scores on the Social Problems and Aggressive behaviours subscales but not on the Rule-Breaking subscale from the CBCL (Gascoigne, et al., 2019). Lastly, one study administered the Social Responsiveness scale, and found deficits on the social communication subscale but not the prosocial behaviour subscale (Stewart et al., 2019a).

Results from meta-analysis revealed that for social behavior the effect in TLE was low with a Hedge’s g of 0.13 (95% CI = −1.26-0.98; Z score = −.24, *p* = 0.80; Table 5). As this range does include zero, the effects were not significantly different from zero, so we cannot reject null hypotheses. Thus, on average, patients were not significantly different from non-clinical control groups in social behavior. However, due to the small number of studies included, the results need to be interpreted cautiously for this construct. The effect was also not significant for patients with frontal epilepsy (Hedge’s g of 0.47 (95% CI = −0.44-1.40; Z score = 1.01, *p* = 0.31).

#### Specific emotions: Patients vs. non-clinical control

We included studies that specifically measured and reported performance for specific emotions, anger, sadness, fear, surprise, happiness, disgust. All patients performed poorly relative to non-clinical group evident by the Hedge’s g and Z values (Table 5). Based on the effect size values, disgust (Hedge’s *g* = 0.42; 95% CI = 0.21-0.59; Z score = 3.78, *p* < 0.001), fear (Hedge’s *g* = 0.40; 95% CI = 0.17-0.59; Z score = 3.32, *p* = 0.001), anger (Hedge’s *g* = 0.36; 95% CI = 0.04-0.61; Z score = 2.23, *p* = 0.025), and sadness (Hedge’s *g* = 0.38; 95% CI = 0.16-0.56; Z score = 3.28, *p* = 0.001) showed large effect sizes, meaning that patients significantly and substantially recognized these emotions more poorly than the non-clinical group.

#### Specific theory of mind task: Patients vs. non-clinical control

For the specific theory of mind tasks, we included Faux Pas, False Belief, and Reading the Mind in the Eye tests as common tasks used in the literature (Table 5). Overall, patients showed poorer performance in all these measures relative to the non-clinical group, with large effect sizes. However, in the Faux Pas test, more studies (14 studies) were included (Hedge’s *g* = 1.15; 95% CI = 0.57-1.72; Z score = 3.94, *p* < 0.001) and False Belief (Hedge’s *g* = 1.84; 95% CI = 0.47-3.21; Z score = 2.64, *p* < 0.001) and RMET (Hedge’s *g* = 0.86; 95% CI = 0.54-1.18; Z score = 5.33, *p* < 0.001) only included 2 and 6 studies, respectively.

#### Quality ratings

The results of quality rating using the 17-item Downs and Black checklist revealed that each study included in the meta-analytic review was deemed to be of low methodological bias. Score for cross sectional studies ranged from 11-16 out of 16 and longitudinal studies’ scores ranged from 12-16 out of 18 points. Overall, the studies reported in the emotion recognition domain were assessed as exhibiting the lowest quality when compared to the other social cognitive domains, albeit still within the range considered to be ‘low risk’. Most studies failed to report the statistical metrics required such as reporting actual probability (item 8) or estimation of random variability in the data (item 6). Only few studies included confounding variables as covariates in the analyses, but all studies scored perfectly for the methodological biases and all studies had sufficient power. The ratings of each study are included in the supplementary tables 4-6.

## Conclusion

We examined the four major domains of social cognition: emotion recognition, theory of mind, empathy, and social behavior, among patients with temporal and frontal lobe epilepsy. Both narrative synthesis and meta-analyses were conducted to compare patients with temporal lobe and frontal lobe epilepsy (TLE and FLE, respectively) with non-clinical controls as well as comparing between left and right-side of lesion among TLE patients. Next, we discuss the theoretical and clinical implications of these findings alongside suggestions to support routine clinical assessment of social cognition for people with epilepsy.

### Magnitude of Social Cognitive Deficits Following Epilepsy

This is the first review, to our knowledge, to comprehensively examine functioning across the four major domains of social cognition in people with TLE and FLE. Compared to non-clinical controls, these focal epilepsy groups exhibited moderate to large deficits across each of the four social cognitive domains. These deficits were most notable within the domains of emotion recognition and theory of mind, which are in line with previous meta-analyses in this area suggesting deficits in the emotion recognition and theory of mind in focal epilepsy patients (Stewart et al., 2016; Edwards et al., 2017). Our findings are also in accordance with the broader literature on social cognitive deficits among many clinical populations (Cotter et al., 2018). Additionally, our results converge with previous meta-analyses suggesting the magnitude of social cognitive impairment varies according to focal epilepsy syndromes. Specifically, FLE patients exhibit significantly greater impairment in emotion recognition compared to TLE patients while the effects were comparable for theory of mind.

More specifically, the summary effect size for emotion perception was larger in patients with FLE compared to non-clinical controls than those with TLE compared to non-clinical controls. This difference is likely to reflect differential disruption to the underlying neural substrates subserving emotion perception according to the location of the epileptogenic focus. Converging evidence suggests importance of the prefrontal cortex and limbic areas during processing emotional facial expression (Todorov 2013; Lopatina et al., 2018). While the importance of limbic areas has been established in the literature for processing social cues (Pessoa 2010; Adolphs, 2008; Adolphs 2010), damage to prefrontal cortex, such as the case of Phineas Gage, has adverse impact on the processing of social and emotional cues (Perry et al., 2017; Martins et al., 2012). Our results are in line with this notion that prefrontal areas are critical for processing and regulation of social and emotional cues (Buhl et al., 2014) and perhaps insults to this area could result in more adverse impact on the performance among patients with FLE relative to TLE. Importantly however, our methodology did not permit direct comparison of FLE and TLE patients across the four domains of social cognition. Hence, further direct comparison is warranted. Additionally, the precision of the findings extracted from studies with FLE patients is uncertain given the small number of eligible studies to date. Additional studies are needed to investigate social cognitive functions in FLE and confirm the current findings.

Additionally, it is worth mentioning that the age of disease onset was early among patients with FLE, relative to the TLE group. The ability to understand emotions may facilitate the acquisition of emotional skills as the child continues to develop (Conte et al., 2019). Thus, insults to these areas could impact the ability to perceive social and emotional cues in later life more severely (Korkmaz 2011). It remains an open question for further investigation, however, to clarify the phenomenology and social cognitive performance, and specifically emotion recognition, among patients with epilepsy at different stages of disease’ development.

### Laterality of Temporal Lobe Epilepsy

Our results indicate that patients with RTLE have greater difficulties in the theory of mind domain compared to those with LTLE. Previous studies on the laterality of social cognitive functions indicate that the right hemisphere may play a more important role in theory of mind than the left hemisphere (Giovagnoli et al., 2011; Bora et al., 2016). A recent meta-analysis on stroke patients also indicated that the magnitude of social cognitive impairment was greater following right than left hemisphere insults (Adam et al., 2019). These hemispheric differences are in keeping with studies showing that the right temporo-parietal junction is particularly important for theory of mind processing (Saxe & Wexler, 2005).

### Relationship Between Task Features and Task Performance

A recent meta-analysis suggests that the temporo-parietal junction, medial prefrontal cortex, posterior superior temporal sulcus, and inferior parietal lobe are critical in theory of mind (Schurz et al., 2014). Given that these areas are disrupted by focal epilepsies such as TLE and FLE, it is not surprising that patients with epilepsy might show difficulties in the performance of theory of mind tasks. Interestingly, our results suggest distinctions in the types of tasks within the construct of theory of mind that are affected by epilepsy. While studies found large effects for Faux Pas, False Belief and Reading the Mind in the Eye tests, the Faux Pas test is more affected in these patients than the other two tasks. It is possible that this difference simply reflects difference in the demands of each task on cognitive and executive functions which is in line with Stewart et al.’s findings (2016). For example, the Faux Pas test relies more heavily on language understanding and skills than the Reading the Mind in the Eye test. While there is an association between language and theory of mind skills (Milligan et al., 2007), this explanation does not seem to be supported by the difference in performance between LTLE and RTLE patents. RTLE patients performed worse than LTLE patients in our analyses, however, none of the studies reported the relationships between executive functioning, and theory of mind skills. A potential avenue for future research is to consider how visual and verbal processing could account for social cognitive performance in this population. See also supplementary table 8 for summary of previous studies and inclusion of executive functioning measures. Additionally, we highlightthe lack of systematic research establishing the psychometric properties of social cognitive measures for use with people with epilepsy. Future studies confirming the psychometric rigour of these measures for this population will enhance the confidence in interpreting the clinical and theoretical significance of results.

### Clinical Recommendation and Future Directions

Accumulating evidence emphasizes that, in addition to the influence of disease characteristics and cognitive or psychiatric comorbidities, changes in social cognition impacts psychosocial outcomes in people with epilepsy (Yogarajah & Mula, 2019). Clinically, deficits in social cognition remain under-recognized, likely due to oversight of this domain during clinical evaluation (Wilson et al., 2015). Deficient social cognition has wide-ranging functional implications across multiple psychosocial domains (Wang et al., 2015). Therefore, disclosure of psychosocial challenges ought to trigger consideration of social cognitive impairment as a possible etiological factor and prompt more targeted clinical and psychometric enquiry. While difficulties in other cognitive (e.g., language or visual processing) or psychiatric domains (e.g., depression) may be the root of dysfunctional social behavior, the modularity of social cognition means that impairment may also occur in isolation (McDonald & Cassel, 2017). Further, the integrity of social cognition cannot be determined from performances across other higher cognitive domains alone, necessitating the inclusion of targeted measures of social cognition within comprehensive psychometric batteries.

In terms of clinical practice recommendations, we encourage clinicians to remain vigilant to indicators of social cognitive dysfunction in patients with TLE and FLE. To guide clinic-based screening of problematic social behaviour, we provide some example probes that clinicians can use to guide their clinical interview (see Table 7). This list is not exhaustive but provides a starting point for clinicians to explore suspected concerns about an individual’s social behavior. Collateral information is vital because deficits in social cognition may be subtle or self-appraisal limited, for example by insight/awareness, emotional state, feelings of stigma, and capacity to understand and articulate responses to questions about abstract social cognitive constructs.

**Table 7.**
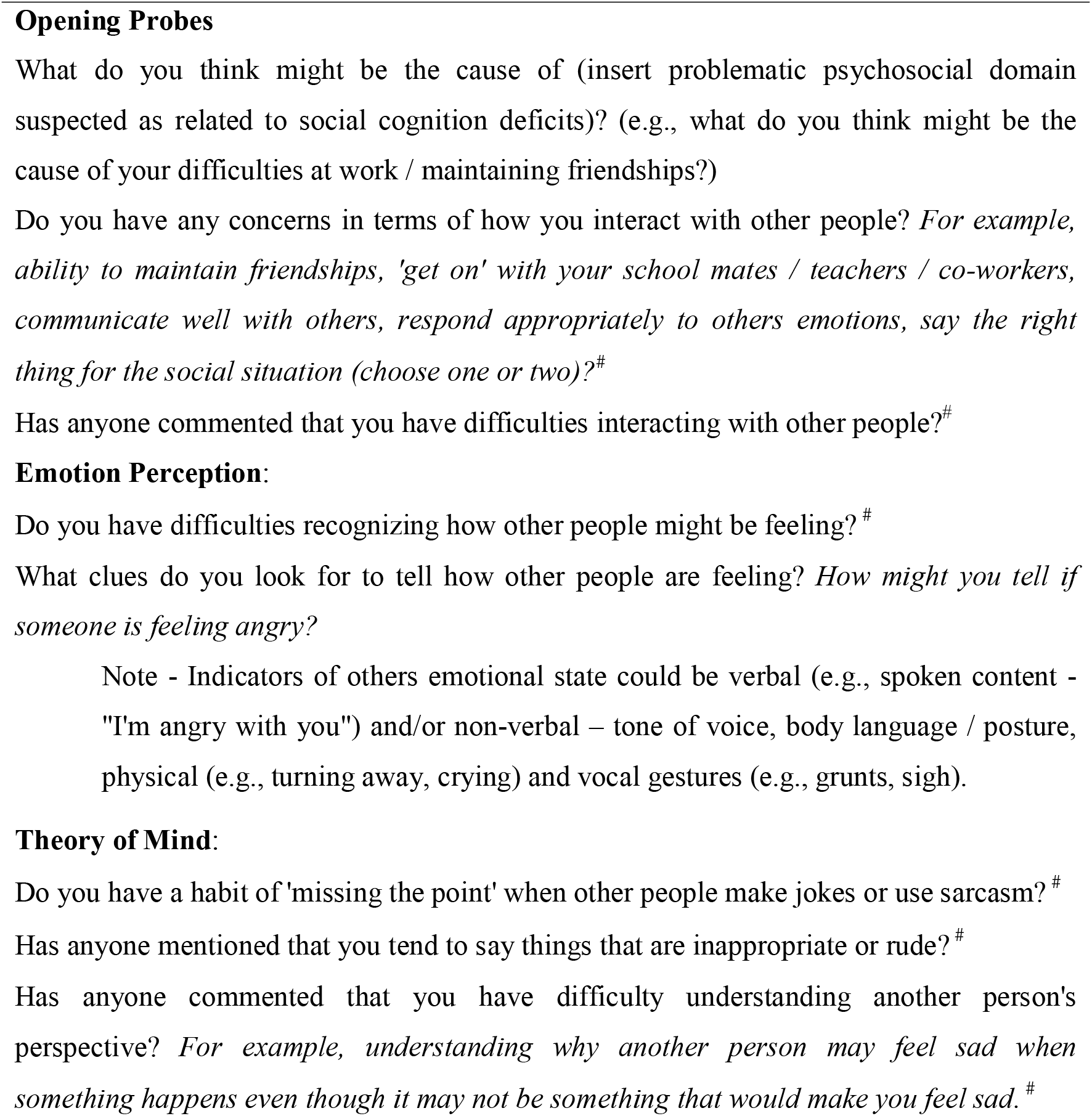
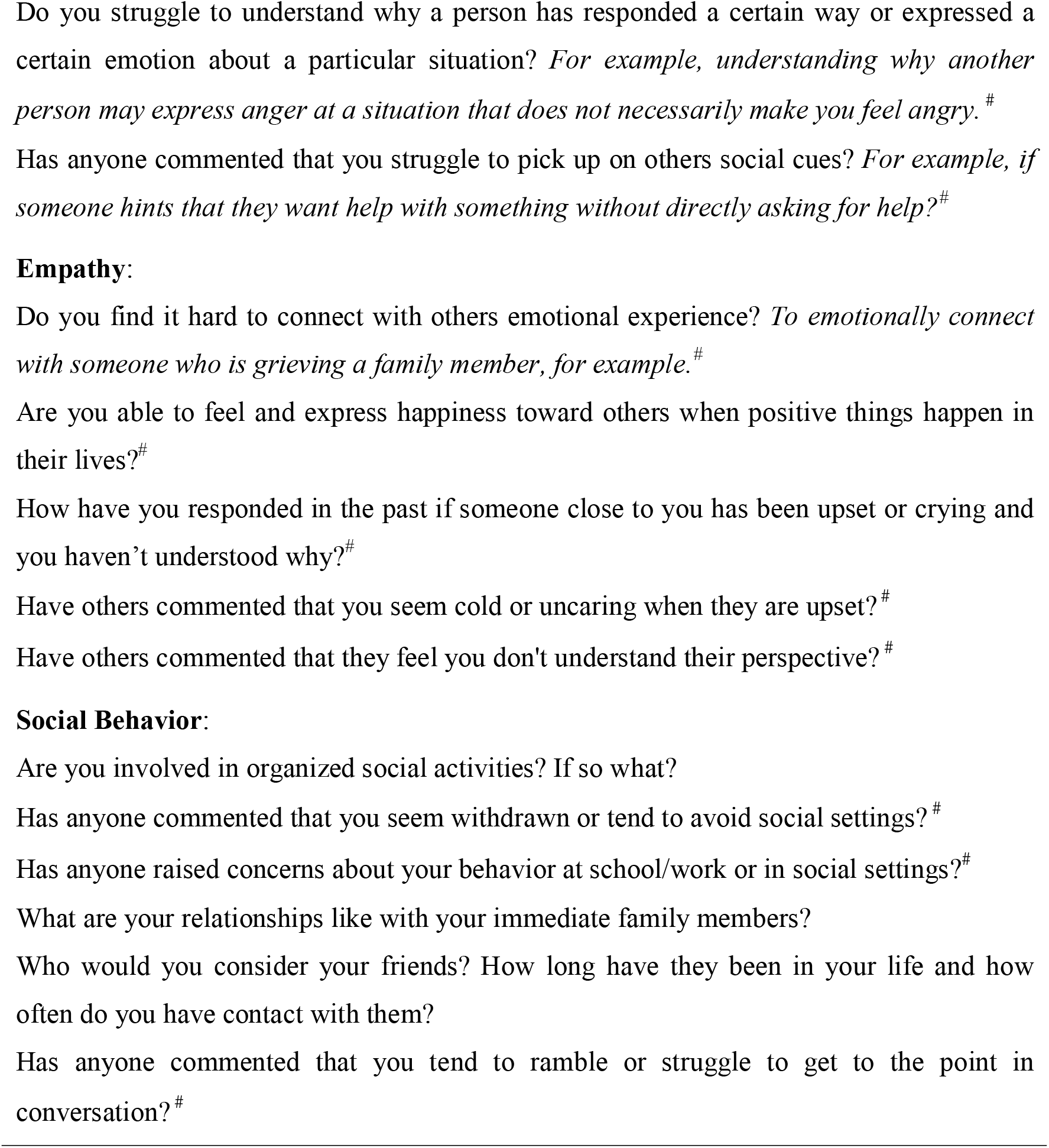
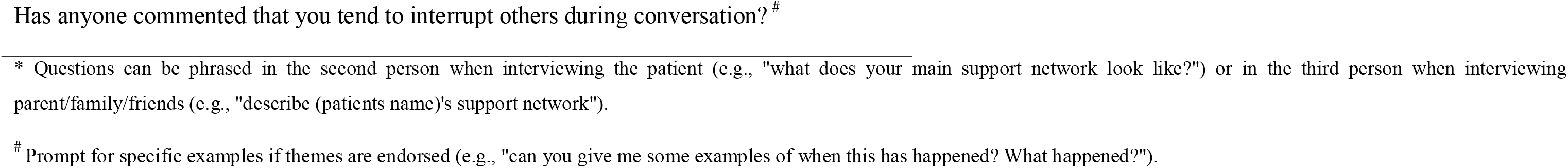
Probe Questions for Social Cognition Domains.

If clinic-based screening further substantiates concerns about dysfunctional social behaviour multidisciplinary psychiatric, psychological and neuropsychological assessment should be requested to comprehensively investigate the aetiological factors at play. Multidisciplinary evaluation is recommended to optimise accurate differential diagnosis given the overlap of personality vulnerabilities, psychopathology and cognitive impairment as drivers of impaired social behaviour. While input from multiple specialties is desirable, we recognise that this may not be feasible in some services due to staffing and/or time constraints. Therefore, in practice, referrals to available specialists may be initiated sequentially according to the balance of evidence obtained during initial screening (e.g., evidence of co-morbid psychopathology may indicate initial referral to psychiatry whereas evidence of co-occuring cognitive difficulties may indicate initial referral to clinical psychology or neuropsychology for psychometric testing). To assist clinical psychologists and neuropsychologists in test selection, we provide tentative suggestions, pending confirmation via systematic review, of measures to evaluate each of the four social cognitive domains in people with epilepsy (see supplementary table 7). While we based our suggestions on the most frequently used measures within this review, a systematic review of the psychometric properties of social cognitive measures pertintent to people with epilepsy is a priority for future research.

Multiple data sources are required to appropriately determine whether social difficulties are primary manifestations of impaired social cognition or secondary to one or more psychosocial comorbidities. Interpretation and formulation of findings from multiple psychometric tests is a core capability of clinical psychologists and neuropsychologists; disciplines which are well placed to contribute to the differential diagnosis of the basis of impaired social behavior.

## Conclusion

This study is the first to investigate all four domains of social cognition in patients with epilepsy using both systematic and statistical meta-analysis reports. We investigated performance of people with TLE and FLE on emotion recognition, theory of mind, empathy and social behavior. Our results from meta-analysis revealed that patients with FLE demonstrated poorer performance compared to those with TLE, mainly in emotion recognition tasks. While our results highlight the variability of social cognitive functions in the population of epilepsy patients, it also demonstrates the importance of the frontal lobe in social cognitive performance. Our results provide future directions for clinical and experimental research with the emphasis on the inclusion of social cognitive tasks in the clinical setting. We also highlight a gap in the literature in studies focussing on the domains of empathy and social behavior in patients with focal epilepsy and psychometric properties of these measures among people with epilepsy. Furthermore, additional experimental research is needed to explore the overlapping cognitive and mental disorders related to social cognition in patients with focal epilepsy. It is of utmost importance for clinicians to consider social cognitive deficits as a potential contributory factor in their assessment of impaired social behavior and which could help to move toward a more holistic treatment plans for this population.

## Data Availability

Data is available upon request from the corresponding author

## Acknowledgement

Authors would like to thank Priscilla Tjokrowijoto for help with literature search. MZ was funded by the UQ Development Fellowship (Research and Teaching) at the time of conducting this review. Data were collected using the New Staff grant money at the Centre for Advanced Imaging (no grant number). Authors declare no conflict of interest.

## Notes

### Competing Interest Statement

The authors have declared no competing interest.

### Funding Statement

This project was funded by RBWH foundation

### Author Declarations

HREC/17/QRBW/146

